# Effect of information about COVID-19 vaccine effectiveness and side effects on behavioural intentions: two online experiments

**DOI:** 10.1101/2021.03.19.21253963

**Authors:** John R. Kerr, Alexandra L. J. Freeman, Theresa M. Marteau, Sander van der Linden

**Affiliations:** Winton Centre for Risk and Evidence Communication, University of Cambridge, Wilberforce Road, CB3 0WA Cambridge, UK; Department of Psychology, School of Biological Sciences, University of Cambridge, Downing Street, CB2 3EB Cambridge, UK; Department of Public Health and Primary Care, University of Cambridge, Worts Causeway, CB1 8RN, Cambridge, UK

**Keywords:** COVID-19, vaccination, hesitancy, communication, vaccine messaging

## Abstract

The success of mass COVID-19 vaccination campaigns rests on widespread uptake. However, although vaccinations provide good protection, they do not offer full immunity and while they likely reduce transmission of the virus to others, the extent of this remains uncertain. This produces a dilemma for communicators who wish to be transparent about benefits and harms and encourage continued caution in vaccinated individuals but not undermine confidence in an important public health measure. In two large pre-registered experimental studies on quota-sampled UK public participants we investigate the effects of providing transparent communication—including uncertainty—about vaccination effectiveness on decision-making. In Study 1 (*n* = 2,097) we report that detailed information about COVID-19 vaccines, including results of clinical trials, does not have a significant impact on beliefs about the efficacy of such vaccines, concerns over side effects, or intentions to receive a vaccine. Study 2 (*n* = 2,217) addressed concerns that highlighting the need to maintain protective behaviours (e.g. social distancing) post-vaccination may lower perceptions of vaccine efficacy and willingness to receive a vaccine. We do not find evidence of this: transparent messages did not significantly reduce perceptions of vaccine efficacy, and in some cases increased perceptions of efficacy. We again report no main effect of messages on intentions to receive a vaccine. The results of both studies suggest that transparently informing people of the limitations of vaccinations does not reduce intentions to be vaccinated but neither does it increase intentions to engage in protective behaviours post-vaccination.

---

At the time of writing more than 2.5 million people are estimated to have died from COVID-19 (WHO, 2021) and wide-ranging restrictions have been placed on people across the globe, with immense social and economic consequences (Chowdhury et al., 2021; Saladino et al., 2020). Mass vaccination has been heralded as a ‘checkmate’ move to end the pandemic (Pearl, 2020). However for lasting and widespread protection, this relies firstly on vaccines being developed that significantly reduce transmission of the disease rather than simple individual protection, and secondly, sufficient uptake of the vaccine to produce ‘herd immunity’—a level of population immunity at which an infectious agent is no longer able to spread (Randolph & Barreiro, 2020). Estimates of the percentage of the population required to be vaccinated to achieve this goal vary (and depend on the local reproduction rate, *R*, of the virus), with most estimates in the 60-80% range (Anderson et al., 2020; Kadkhoda, 2021).

A sizable minority of people around the world express hesitancy towards taking COVID-19 vaccines (C. Lin et al., 2021)—defined as “delay in acceptance or refusal of vaccines despite availability of vaccination services.” (WHO, 2014, p. 7). Factors such as religion, gender, political ideology, and trust in medical and scientific institutions have been shown to be associated with vaccine hesitancy, both in general and regarding COVID-19 vaccines specifically (de Figueiredo et al., 2020; Kerr et al., 2020; C. Lin et al., 2021). While these broader factors are important, research has also shown that specific beliefs about and attitudes towards COVID-19 vaccinations are also closely linked to vaccination intentions (Freeman et al., 2021; C. Lin et al., 2021). Whether these attitudes can be, and should be, changed by communication ‘campaigns’ is a matter of active debate worldwide.

## Transparent communication of vaccine risks and benefits

Many public health commentators and researchers have called for transparent communication of COVID-19 vaccine information, including the efficacy and side effects reported from clinical trials, to improve vaccine uptake (Cohen et al., 2020; Nature, 2020; Quinn et al., 2021). Some, for example, Rzymski et al. (2021), think that such information may change individuals’ vaccine beliefs: “The general public must be given access to the pivotal information on the authorized vaccines, and that their approval is based on the evidenced benefits that outweigh the potential risks of vaccine administration” (p. 3). Others see such transparency as more a matter of ensuring public trust in science, or simply an ethical imperative (Blastland et al., 2020). Such calls demand empirical evidence of their potential effects, and hence are the impetus for the current study. What are the effects of such communications on individuals’ beliefs about COVID-19 and their intentions to receive a vaccine? We note three important considerations in approaching this question.

Firstly, vaccination is an invasive medical procedure (Cousins et al., 2019) with well-defined (if small) risks to the individual. As such, in some countries it is treated as an individual medical decision and requires informed consent (in others it is treated as a public health policy and can be mandated without the option of consent or refusal – for some discussion see O’Neill, 2020; O. O’Neill, 2017; Parmet, 2005; Reiss & Karako-Eyal, 2019). In the case of the COVID-19 vaccines, until recently there was no evidence that they reduced transmission of the disease to others and therefore were more accurately considered a method of individual rather than public protection. Many might argue, therefore, that clearly and transparently communicating what is currently known about COVID-19 vaccines should be considered important from an ethical (GMC, 2021) (and potentially legal; Marco-Franco et al., 2021) standpoint, regardless of any expected effect of such messages on individuals’ vaccination intentions.

Second, it is currently unclear that transparent communication of COVID-19 vaccine information would necessarily affect vaccination rates in any predictable way. On the one hand, explaining the results of clinical trials and the regulatory procedures in place may serve to alleviate concerns over vaccine efficacy and safety and ultimately increase intention to vaccinate as suggested by some researchers (Freeman et al., 2021). On the other, as noted by Petersen et al. (2020), it is also possible that individuals may find the information *more* concerning, increasing their perception of vaccines as relatively more dangerous or ineffective. That is, some people may find the reported frequency of COVID-19 vaccine side effects to be higher, or efficacy of vaccines to be lower than they expected. Indeed, some (pre-COVID) vaccination messaging studies have reported ‘backfire’ effects, such that well-meaning efforts to increase vaccination intentions actually increase safety concerns and lower vaccination intentions (Rossen et al., 2016).

Third, the notion that COVID-19 vaccine hesitancy is the product of a lack of information reflects a ‘deficit model’ approach to addressing the overarching issue. That is, it assumes simply providing information will lead to attitudes and behaviours in line with the mainstream scientific position (Simis et al., 2016). In the context of vaccination attitudes, Kitta and Goldberg (Kitta & Goldberg, 2017) position the deficit model as “…the presumption that vaccination opposition and/or refusals are primarily driven by insufficient understanding of the facts regarding vaccination risks and benefits.” (p. 2). Within science and health communication fields the deficit model approach has received widespread criticism as ignoring the myriad social, cultural and psychological factors that drive risk perceptions and attitudes toward medical and scientific issues and technologies (Kitta & Goldberg, 2017). However, COVID-19 vaccines are a very new medical intervention and a lack of information may play a greater role in attitudes and intentions compared to resistance towards other, more established vaccines. This point is explicitly noted by Opel et al. (2020) in comparing general vaccine hesitancy to COVID-19 vaccine hesitancy. They write: “The former is steeped in a complex sociocultural milieu of consumerism, pervasive misinformation, and the rejection of objective, scientific evidence as truth. The latter may simply represent a deficit of the data needed to make an informed decision.” (p. 1)

Regardless of the considerations outlined above, public health authorities around the world *are* currently communicating information about COVID-19 vaccines to citizens. Thus we conducted these experiments with the intention of generating empirical data on the effects of such real-world communications, in order to inform debates about the best kinds of communication to serve the intentions of different communicators, whether they be attempting to inform or persuade.

## Beliefs about COVID vaccines and vaccine hesitancy

Numerous cross-sectional surveys around the world have identified perceptions of COVID-19 vaccines as safe and effective as key predictors of COVID-19 vaccination intentions (e.g. Callaghan et al., 2021; Freeman et al., 2021; Karlsson et al., 2021; Y. Lin et al., 2020; Paul et al., 2021; Wang et al., 2021; Wong et al., 2020). Indeed, a recent systematic review of such studies (C. Lin et al., 2021) concludes that perceptions of vaccine safety and effectiveness are ‘universally’ consistent determinants of COVID-19 vaccine hesitancy (p. 8). Further, surveys which specifically asked COVID-19 vaccine hesitant participants their reason for vaccine refusal or delay consistently report that concerns over safety and efficacy are among the most common reasons given (Callaghan et al., 2021; Fisher et al., 2020; Frank & Arim, 2020; Ruiz & Bell, 2021; Yigit et al., 2021; Yoda & Katsuyama, 2021). Similarly, qualitative research has also highlighted these concerns as reported drivers of COVID-19 vaccine hesitancy among minority and at-risk groups (Lockyer et al., 2020; Momplaisir et al., 2021; Williams et al., 2020). Such concerns may be fuelled by COVID-19 vaccine misinformation (Hotez, 2020) and recent research has identified susceptibility to misinformation as a correlate of COVID-19 vaccine hesitancy (Roozenbeek et al., 2020).

Such findings align with Health Belief Model theoretical framework which positions the perceived risks and benefits of a given health behaviour as key predictors of intentions to engage in that behaviour (alongside perceptions of disease threat; Carpenter, 2010; Maiman & Becker, 1974). The Health Belief Model has informed previous vaccine communication research. For example, Jones et al. (2015) show that exposure to vaccination campaign advertisements is associated with increased willingness to receive a H1N1 vaccine, and that this effect is partially mediated by belief that the vaccine will prevent disease and belief that the vaccine would not have a negative impact on one’s health. The authors conclude that addressing perceived barriers (e.g. perceptions of vaccine risk) may be the most effective target for interventions to improve vaccine uptake.

## The effect of COVID-19 vaccine communications on intentions

Few studies have experimentally investigated the effects of COVID-19 vaccine information on related attitudes and intentions, and most have focused on hypothetical vaccines due to the lack of approved vaccines at the time of research. In discrete choice-type experiments, participants unsurprisingly express greater preference for more effective (hypothetical) vaccines with fewer side effects, *ceteris paribus* (Borriello et al., 2021; Kaplan & Milstein, 2021; Kreps et al., 2020; Motta, 2021; Schwarzinger et al., 2021). However, experimental studies which have examined the impact of COVID-19 vaccine information on behavioural intentions report mixed results. For example Petersen et al. (2020) report that transparent positive messages heralding a safe and effective fictional COVID-19 vaccine (70% efficacy with a 1 in 10,000 incidence of serious side effects) decreased vaccine scepticism relative to vague or negative transparent messages. However, no control group was included in the study—leaving it unclear which interventions shifted beliefs relative to a baseline. In another messaging experiment, Motta et al. (2021) report that messages framed to emphasise the collective benefits of vaccination or the risks of not vaccinating increased participants’ vaccine intentions—however the presence or absence of information about the rigorousness of clinical trials had no impact on intentions. Palm et al. (2021) find that messages highlighting vaccine efficacy and safety had a marginally significant effect on vaccine intentions relative to a control group, while an opposite message claiming planned vaccines were neither effective nor safe had no main effect on intentions. Conversely, in a pre-post design experiment Loomba et al. (2021) report that exposure to a range of factual information about potential COVID-19 vaccines has no effect on vaccine intentions, while COVID-19 vaccine misinformation decreases vaccination intentions. Duquette (2020) reports that short messages framing vaccination as protecting either oneself or others have no effect on reported vaccination intentions (but notes that there are some experimental effects among non-white participants).

Notably, all of the studies reviewed above focus on fictional or hypothetical vaccines, and none examine the impact of information on beliefs about COVID-19 vaccines—such as perceptions of efficacy or safety—separately from intentions to receive a vaccine. We are aware of only one study which has used actual results from clinical trials as part of an experimental intervention. Altay et al. (2021) paid French participants to spend 10 minutes interacting with an extensive Q&A interface in which users could select questions and read answers (the complete Q&A comprised more than 20 pages of text). Some of the answers included preliminary results from clinical trials outlining vaccine efficacy (95% for Pfizer/BioNTech and Moderna, and <70% for AstraZeneca) and noted some side effects without giving any indication of frequency. Altay et al. report that participants who interacted with the Q&A were more likely to report intentions to receive a COVID-19 vaccine when available and expressed more positive attitudes towards vaccines, compared to a control group. However, it is unknown how many participants actually accessed or read the information on vaccine safety and efficacy.

In Study 1, we examine the effects of reading comprehensive information from recent clinical trials (in two different formats) on individuals’ intentions to receive a vaccine. In addition to investigating vaccination intentions we also include several outcome measures examining: participants’ belief that COVID-19 vaccines are effective, concern over side effects and concern over speed of regulatory approval (using the Oxford COVID-19 Vaccine Beliefs Scale; Freeman et al., 2020). We also include two further exploratory messages as experimental conditions. The first focuses on addressing concerns over the speed of vaccine development and approval, which is also associated with vaccine hesitancy (Freeman et al., 2021) and the second presents a mechanistic description of how vaccines, including the new mRNA vaccines, work. Mechanistic explanations have been shown to reduce risk perceptions and shift attitudes in other scientific domains (McPhetres et al., 2019; Ranney & Clark, 2016).

## Post-vaccination protective behaviour messaging and vaccine confidence

Although the reported efficacy of some vaccines in preventing SARS-CoV-2 symptoms is very high (e.g. 94.1% for the Moderna mRNA vaccine; Baden et al., 2021), there is considerable uncertainty about the effectiveness of others, particularly in some groups of people (e.g. AstraZeneca in over-65s; Torjesen, 2021). At the time of writing, the extent to which vaccines will prevent transmission of the virus is even more unclear and the majority of people in most countries are unvaccinated (The Lancet Microbe, 2021). This has lead public health authorities to emphasise the need for vaccinated individuals to continue engaging to preventative behaviours such as social distancing, frequent handwashing and wearing a mask in public spaces to prevent the spread of the virus (CDC, 2021; NHS, 2021; Su et al., 2021).

Do such messages increase intention to continue protective behaviours post-vaccination? And do they have unintended consequences? It is possible that such messaging could lead individuals to think that vaccines are ineffective, and/or reduce intentions to receive a vaccine as some perceived benefits (e.g. not having to wear a mask or social distance) are no longer present. As Savulescu (2021) speculates, freedom from social distancing and mask wearing requirements may be a powerful personal incentive for some individuals to receive a COVID-19 vaccine (see also Prasad, 2021). Equally, could health messages in use to encourage the uptake of the vaccine which minimise uncertainty and risks (e.g. ‘vaccines are safe and effective’) encourage vaccinated people to stop engaging in protective behaviours once vaccinated?

In Study 2 we test these possibilities, providing participants with messages encouraging vaccination, with or without caveats about effectiveness and additional reminders of the need to maintain protective behaviours once vaccinated.

### Study 1

This study sought to answer the following research question: does information specifically targeted to beliefs about vaccine efficacy, or concerns about safety shift these beliefs, and, if so, do these changes in turn have an impact on COVID-19 vaccination intentions? We note that the different messages presented here were drawn from publicly available information and do not vary systematically in content, thus our main focus is comparing the different messages with the control group. We would also add that none of the messages were directly persuasive regarding vaccination, that is, none explicitly stated that the reader *should* accept a vaccine if offered. This study was pre-registered (https://aspredicted.org/blind.php?x=we7am9) and ethical oversight was provided by the Psychology Research Ethics Committee at the University of Cambridge (PRE.2020.034 with amendment on 9^th^ January 2021).

## Methods

### Participants

An a priori power calculation performed using GPOWER (Erdfelder et al., 1996) indicated a total sample size of at least N = 1,865 would be required to detect an effect size of f = 0.1 in a one-way ANOVA with five groups, at 95% power and an alpha level of .05. We deliberately recruited a larger sample to account for the exclusion criteria detailed below.

Adult participants resident in the U.K. were recruited through two online panels: Respondi (Respondi.com) and Prolific (prolific.co), between 13-17 January 2021. Participants recruited via Respondi were quota sampled (using Qualtrics survey software) to match the age and sex distribution of the UK population. Participants recruited via Prolific were quota sampled by the platform to match the UK population in terms of age, sex, and ethnicity (Prolific, n.d.).

A total of 2,488 participants completed the survey (Respondi n = 1,233; Prolific n = 1,255). As per our pre-registration, 391 participants were excluded for the following reasons: failing an instructional attention check (‘Please select ‘somewhat agree’…’, n = 279), reporting having already received a COVID-19 vaccine (n= 90), providing an age under 18 (n = 2) or higher than 100 (n = 3).

This resulted in a final sample for analysis of 2,097 (*M*_Age_ = 43.2, *SD* = 15.5; 51.7% female; median highest education: Bachelors degree or equivalent; 87% White ethnicity).

### Materials and methods

After providing consent, participants answered questions about their attitudes and beliefs around COVID-19 before being randomised via the Qualtrics randomisation function to an experimental group and answer questions about their COVID-19 vaccine beliefs and intentions. They were blinded to the randomisation. All participants then completed a section of demographic questions.

#### Information Texts

There were four different information conditions and one control condition in the experiment. Participants in the control condition were presented with no information, but instead proceeded directly to post-information items. Participants in all other conditions were presented with information on a single webpage within the survey. All started with the following text: “Several new vaccines have been developed to protect people against COVID-19, which is the disease caused by the SARS-Cov2 coronavirus. These vaccines have been developed and tested throughout 2020 and are now becoming available to the public.”

The content of each text is briefly summarised in Table 1; the full text of each condition can be found in the supplementary information (Appendix 1). The **Factbox** format is a tabular format allowing people to compare potential benefits and harms in those taking and not taking a particular medical treatment. It has been shown to help people understand and remember such information (Brick et al., 2020; McDowell et al., 2016; Schwartz & Woloshin, 2013). The text **Q&A** format was an edited version of text from the European Medicines Agency website. Both were designed to address concerns about efficacy and side effects, but the Factbox condition was much more explicit about side effects. Data reported in the Factbox and Q&A conditions was based on a trial of the Moderna vaccine, but the name was omitted from the text. No source for the information was given. The regulatory **Approval** message condition gave participants information taken from the European Medicines Agency website specifically designed to address concerns about the accelerated approval process of the COVID-19 vaccines. The scientific **Mechanism** message condition gave participants information about how vaccines, and in particular the mRNA vaccines developed against COVID-19, work, designed to address misinformation and misunderstandings about the mechanism and contents of the vaccines, given previous findings on the effect of a meaningful mechanistic explanation(Schneider et al., in preperation).

**Table 1:** Overview of message content in experimental conditions

| Condition | n | Content | Source | Length in words (incl. tables) |
| --- | --- | --- | --- | --- |
| Control | 412 | None | - | - |
| Factbox | 409 | Tables detailing incidence of COVID-19 and side effects in the vaccine and placebo arms of a large clinical trial, both as absolute numbers and percentages. Data was presented separately for trial participants aged 18-65 and aged 65+. A separate table reported rare side effects. | Adapted from US FDA (2020) briefing document | 541 |
| Q&A | 417 | A Q&A format outlining the results of a clinical trial, including the absolute numbers of symptomatic COVID-19 cases and vaccine efficacy. The text also noted several recommendations around the vaccine and known side effects with approximate frequencies, ranging from mild (e.g. pain at injection site, 1 in 10) to severe (facial swelling and facial paralysis, 1 in 1000). | European Medicines Agency (2020a) webpage | 708 |
| Approval | 443 | An overview of the standard and expedited COVID-19 vaccine review processes, highlighting the point that regulatory assessments of COVID-19 vaccine data were undertaken *during* the research and development process, as opposed *after*. A figure depicted this visually. | European Medicines Agency (2020b) webpage | 624 |
| Mechanism | 416 | A description of how vaccines induce immunity and in particular the mechanism by which mRNA vaccines produce antigens, noting the benefits of using mRNA, such scaling up vaccine production quickly. Two figures were included. | Extracts from US CDC (2020) and British Society for Immunology (n.d.) and a figure from Pfizer | 686 |

As the information presented in the conditions was taken from different sources, the content and format varied widely. In particular, we note that, although based on the same trial, efficacy figures in the Factbox and Q&A conditions differed slightly. In the Factbox condition, efficacy was reported separately for trial participants aged 18-65 and 65 and older (95.5% and 86.2% respectively) while the Q&A reported an overall efficacy of 94.1%. The Q&A text also only gave a broad account of side effects, e.g. that mild and moderate side effects “affected more than 1 in 10 people”. This can be compared with the specific side effect frequencies reported in the Factbox text, including the relatively high rate of reported pain at the injection site among vaccinated individuals (e.g. 18.8% in the placebo group and 90.1% in the vaccine group, among 18-64 year olds).

#### Primary Outcomes

Participants completed the seven item Oxford COVID-19 Vaccine Hesitancy Scale, which aims to “assess expressed intent to accept a COVID-19 vaccine” (Freeman et al., 2021, p. 2; example item: *I would describe my attitude towards receiving a COVID-19 vaccine as*; response scale: 1 = *Pretty keen*, 5 = *Against it*; α = .97). The scale was reversed such that higher values indicate a greater willingness or intention to receive a vaccine.

Participants also completed an adapted version of the Oxford COVID-19 Vaccine Beliefs Scale (Freeman et al., 2021) including the subscales capturing: belief that COVID-19 vaccines are effective (*The COVID-19 vaccine is likely to*; 1= *Definitely work for me*, 5 = *Definitely not work for me*; α = .87), concern over COVID-19 side-effects (*The side effects for people of getting the COVID-19 vaccine will be*; 1= *None*, 5 = *Life threatening*; α = .76), and concern about the speed of vaccine development and approval (*The speed of developing and testing the vaccine means it will be*; 1 = *Really safe*, 5 = *Really unsafe*; α = .79). Scores on the efficacy scale were reversed such that higher values indicate a perception that COVID-19 vaccines are effective. The original Freeman et al. (2020) scale dimensions were based on exploratory and confirmatory factor analyses of a larger pool of items. Results from these original analyses indicated some scale items which only loaded weakly onto the identified factors (e.g. a risk question about likelihood of infection had a loading of .45 on the efficacy factor). In light of this, we added one or two face-valid items to each scale and several items were removed (see Appendix 2 for a full description of changes). As expected, the adapted subscales displayed higher reliability than the original scales (however, analyses using the original scale items produced essentially identical results; see Appendix 2).

#### Secondary Outcomes

Participants also completed a second, binary measure of vaccination intention, responding *Yes* or *No* to the question: ‘*If you were offered a COVID-19 vaccine would you get vaccinated yourself?*’ All participants completed two measures adapted from the Decisional Conflict Scale (O’Connor, 1995), with all items prefaced with the stem: ‘*If you were given the option of receiving a COVID-19 vaccine or not today how would you feel about the decision?*’. The first was a three-item measure of feeling informed regarding the decision to vaccinate (example: *I would feel I had made an informed choice;* 1 = *Strongly disagree*, 5 = S*trongly agree*; α = .79). The second was a three-item measure of decision certainty (*I feel sure about what to choose*; 1 = *Strongly disagree*, 5 = S*trongly agree*; α = .92)^1^.

All participants also were asked to estimate the frequency of mild to moderate and severe side effects of COVID-19 vaccines (per 10,000 people vaccinated; this denominator was chosen to align with the figures reported in the Factbox and Q&A conditions). Participants then provided an estimate of efficacy of the ‘most effective COVID-19 vaccine available’ (*By how much do you think the most effective COVID-19 vaccine available reduces the chance of someone becoming ill with the disease?*). Participants entered their estimate on a sliding scale with anchors ‘0% -Doesn’t make any difference’ and ‘100% - Stops all COVID-19 disease’. We refer to this variable as ‘estimated efficacy’ to distinguish it from the perceived efficacy primary outcome,

Participants in the information conditions (i.e. all except control) also completed several measures assessing their perceptions of the information they had just read. These were measures of: how trustworthy (three items, α = .96), engaging (three items, α = .83), and believable they found the information; how much they felt they understood the information; the level of effort required to read the information; and the perceived quality of the evidence underlying the information.

## Results

Participants, on average, reported the messages to be relatively trustworthy (*M*s 5.20-5.61, *SD*s 1.16-1.33), engaging (*M*s 5.17-5.68, *SD*s 1.28-1.36) and believable (*M*s 5.32-5.63, *SD*s 1.21-1.37), with mean scores on these measures above the scale mid-point (range 1-7) across all message conditions (see Appendix 3 for comparisons between conditions).

### Primary outcomes

Reading detailed information about the risks and benefits of vaccination, the vaccine approval process, or how vaccines induce immunity had no significant impact on the main dependent variables: one-way ANOVAs revealed no significant effect of experimental condition on willingness to receive a COVID-19 vaccine, *F*(4, 2091) = 0.72, *p* = .58; belief that COVID-19 vaccines are effective, *F*(4, 2092) = 0.63, *p* = .64, concern over COVID-19 vaccine side effects, *F*(4, 2091) = 1.51, *p* = .20, or concern over the speed of COVID vaccine regulatory approval, *F*(4, 2090) = 0.71, *p* = .58. Results are shown graphically in Figure 1, and descriptive statistics for all outcomes are reported in Appendix 3.

**Figure 1:**
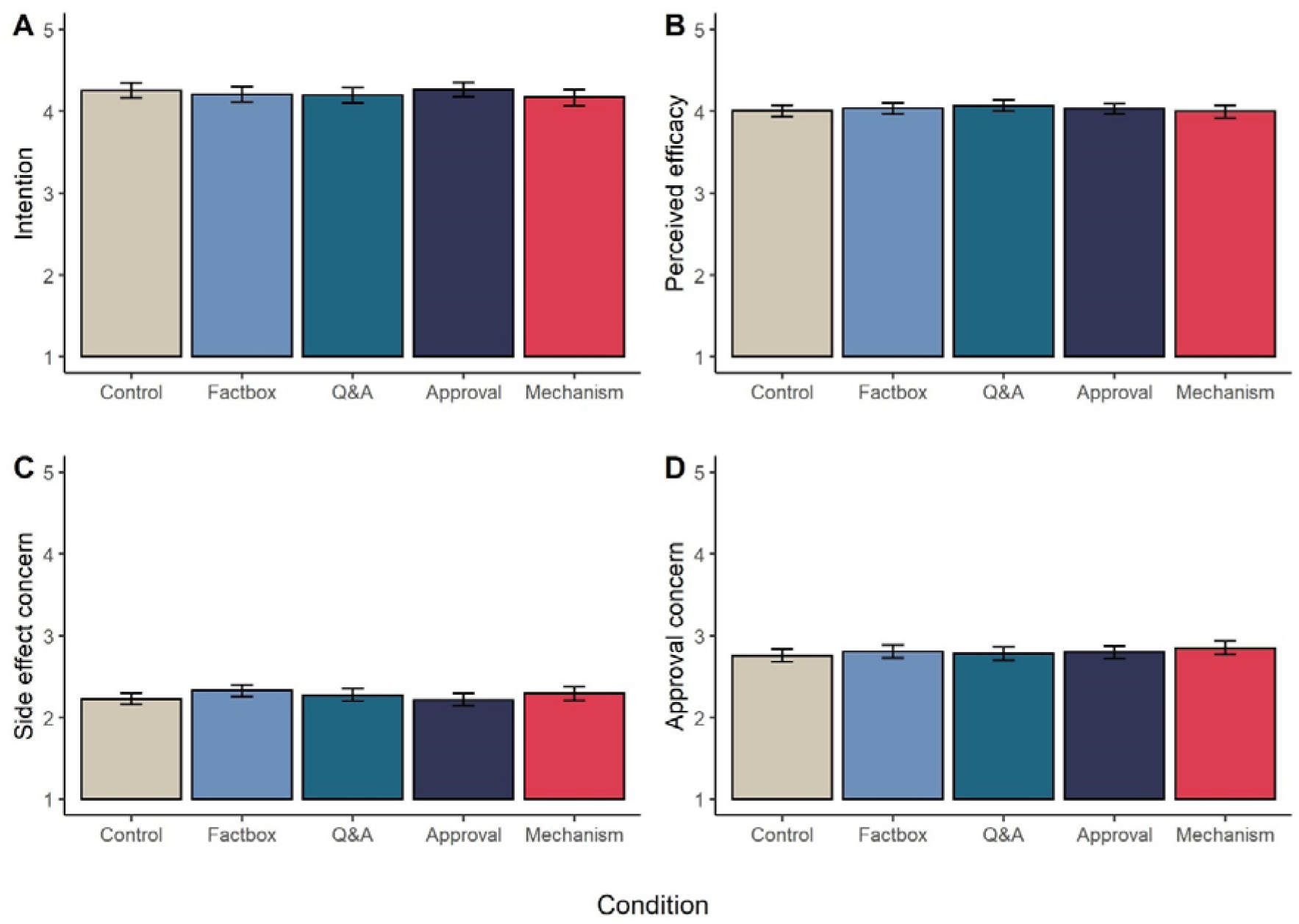
Mean levels (95%CI) of (A) intention to receive a COVID-19 vaccine, (B) perceived efficacy of COVID-19 vaccines, (C) concern over COVID-19 side effects, and (D) concern over speed of COVID-19 vaccine approval process, across the five experimental conditions.

Given the absence of statistically significant effects, we also conducted non-preregistered exploratory Bayesian ANOVAs to investigate the extent to which the data supports a null model (no effect of condition) over a main effect model in which condition does have an effect on the outcome variables. Bayes factors from comparison of models analysing vaccine intentions (*BF* = 1013.40), perceived efficacy (*BF* = 1199.42), concern over side effects (*BF* = 223.48), and concern over speed of approval (*BF* = 1026.08) all indicated ‘very strong’ or ‘decisive’ evidence for the null model over the main effects model (Jarosz & Wiley, 2014; Quintana & Williams, 2018).

### Secondary outcomes

We also asked participants directly if they would receive a COVID-19 vaccine if offered (yes/no). In the control group, 86.9% of participants responded ‘yes’. A chi-squared test of independence indicated that the proportion of ‘yes’ responses did not differ significantly across message conditions (*X*^2^(4)= 5.21, *p* = .27; Factbox: 84.4%; Q&A: 84.7%; Approval: 87.6%; Mechanism: 82.9%).

While the mean level of side effect concern (measured by the adapted subscale of the Oxford COVID Vaccine Beliefs Scale) did not differ between groups, quantitative estimates of mild to moderate side effect frequency (per 10,000 vaccinations) differed between groups, *F*(4, 2080) = 11.60, *p* < .001, η^2^= 0.02. Post hoc tests indicated that the average estimate in the Factbox condition (*M* = 2663.86, *SD* = 2922.01) was significantly higher than that in the control (*M* = 1782.92, *SD* = 2515.07), Q&A (*M* = 1518.20, *SD* = 2313.52), Approval (*M* = 1703.54, *SD* = 2638.12), and Mechanism conditions (*M* = 1919.73, *SD* = 2728.63; all *p* < .001, *d*s .26-.43). There was no main effect of condition on estimates of severe side effect frequency, *F*(4, 2083) = 1.09, *p* = .36^2^, or estimates of vaccine efficacy (i.e. estimated percent of COVID-19 cases prevented): F(4, 2089) = 1.78, *p* = .13 (across conditions, *M*s 74.53-78.18%).

Lastly, we report a significant main effect of condition on responses to the ‘informed’ measure adapted from the Decisional Conflict Scale, *F*(4, 2091) = 10.17, *p* < .001, η^2^ = 0.02 (see Figure 2). Post-hoc analyses revealed that participants in the Factbox condition (*M* = 4.21, *SD* = 0.84) reported, on average, feeling more informed regarding their vaccination decision that those in the control condition (*M* = 3.94, *SD* = 0.91, *p* < .001, *d* = .30) and Mechanism condition (*M* = 4.04, *SD* = 0.89, *p* = .04, *d* = .20. Participants in the Q&A condition (*M* = 4.28, *SD* = 0.86) also reported feeling more informed than those in the control (*p* < .001, *d* = .38), Mechanism (*p* < .001, *d* = .28), and Approval conditions (*M* = 4.05, *SD* = 0.86, *p* = .001, *d* = .27). There was no effect of condition on decision certainty, *F*(4, 2092) = 0.53, *p* = .71.

**Figure 2:**
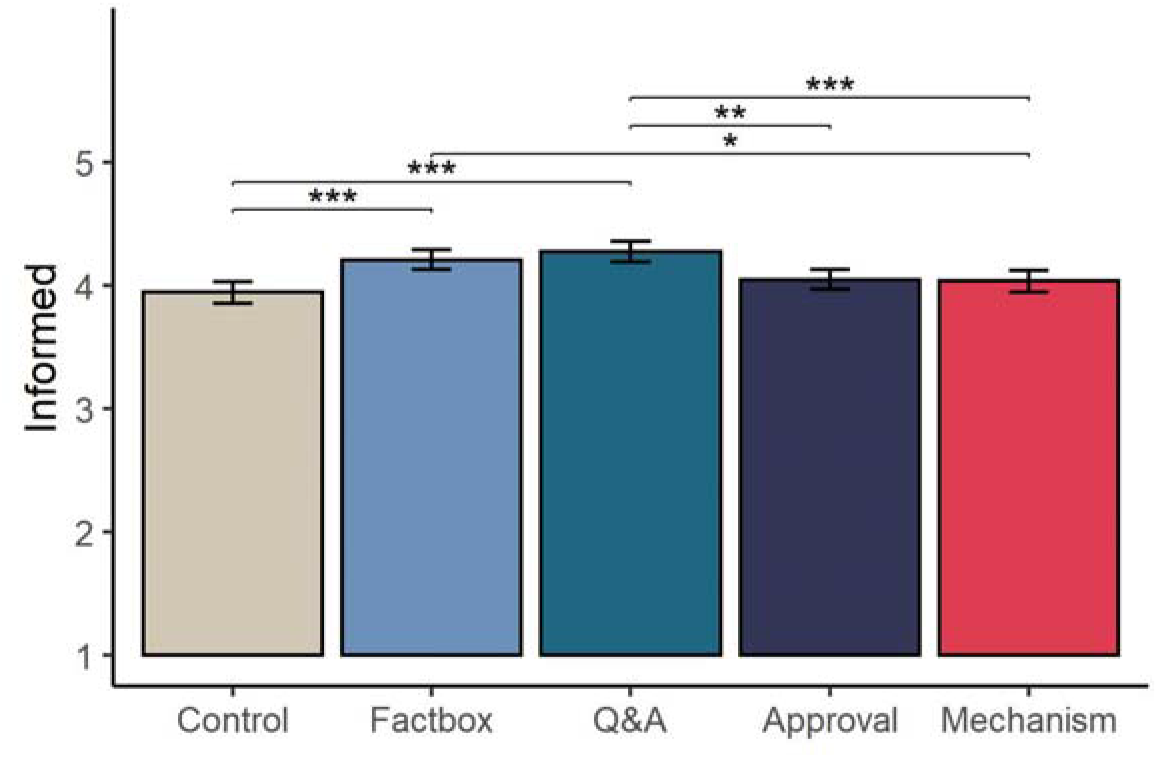
Mean (95%CI) reported level of feeling informed regarding vaccination decision across experimental conditions. Brackets and asterisks indicate significant pairwise differences. \**p* < .05, \*\**p* < .01, \*\*\**p* < .001.

Due to space, results of analyses focusing on ratings of the information (e.g. trustworthiness, believability, ease of understanding) are reported in the supplementary material (Appendix 3).

#### Interim discussion

Detailed information on any of: the efficacy and side effects of a COVID-19 vaccine (in Factbox or Q&A format), regulatory process, or mechanism of vaccine action did not have any impact on vaccine beliefs or intentions. None of the messages in our experiment resulted in a shift (relative to control) in perceptions of efficacy, concerns over side-effects, concerns over the speed of regulatory approval or willingness to receive a vaccine.

The Factbox message, which stated the incidence of a number of side effects in a large clinical trial, did lead participants to give higher estimates of mild side effect frequency compared to the other conditions. Despite this increase, there was no corresponding increase in concern, suggesting that participants did not place much weight on mild side effects when it comes to weighing up the potential harms and benefits of vaccination against COVID-19. Such a conclusion aligns with the findings of a recent conjoint analysis of the effects of hypothetical COVID-19 vaccine attributes on acceptance. Kaplan and Milstein (2021) report that varying rates of minor side-effects (between 50% and 90% likelihood) had no impact on participants willingness to receive a vaccine.

We note that the Factbox and the Q&A messages did not have a significant impact on perceived efficacy. This is despite both messages detailing a high level of vaccine efficacy (95.5% and 94.1% respectively), substantially higher than participants’ average estimated level of efficacy as a percentage of cases prevented (across conditions, *M*s 75-78%). While this might suggest that people were not convinced by the information presented it may well also be the case that participants were adjusting their estimates of efficacy according to the quality of the evidence as they perceived it. The question did not ask ‘what level of efficacy did the information provided say the vaccine has?’, and so it is possible they indicating how well they thought the vaccines would actually perform in the real world. Such levels of adjustment of efficacy according to perceived quality of evidence have been seen in similar experiments around COVID-19 mitigations (Schneider et al., in preperation).

While none of the information had a significant effect on vaccine intentions, some messages increased participants reported awareness of the risks and benefits. Participants who read the Factbox or Q&A materials scored higher on our subjective measure of being informed (i.e. reporting knowing the risks and benefits and feeling informed regarding their vaccination decision) relative to those in the control group. From an ethical standpoint, this improvement in understanding of the potential benefits and harms of vaccination is important, signalling a move towards better informed decision-making on behalf of the participants.

### Study 2

While Study 1 focussed on the effect of messages about COVID-19 vaccines on vaccination attitudes and intentions, Study 2 focussed on a key question regarding the impact of uncertainty over vaccine efficacy and the need for vaccinated individuals to continue engaging in protective behaviours on both vaccine attitudes and intentions to carry out protective behaviour after vaccination (Su et al., 2021).

Specifically, the study addressed two research questions: a) do messages that explicitly caveat the efficacy of vaccines (for self and others) affect perceptions of vaccine efficacy and vaccine intentions, and b) do they have an impact of intentions to follow recommendations post vaccination?

Six different messages were developed, modelled on existing communications from UK public health bodies, designed to vary greatly in the emphasis they put on uncertainty about the efficacy of vaccines and the need to continue protective behaviour once vaccinated. Our aim in presenting extremes of variation in emphasis was to explore the extent to which such messages could influence attitudes and behavioural intentions in an online survey experiment, rather than to identify the effects of particular aspects of such messaging (e.g. the potential protection of self versus others). This study was pre-registered (https://aspredicted.org/blind.php?x=r8wp28) and ethical oversight was provided by the Psychology Research Ethics Committee at the University of Cambridge PRE.2020.034 with amendment on 25^th^ January 2021).

## Methods

### Participants

We planned to compare two sets of messages (long and short; three of each, detailed below) with a control group separately. An a priori power calculation performed using GPOWER (Erdfelder et al., 1996) indicated a total sample size of at least *N* = 1,724 would be required to detect an effect size of *f* = 0.1 in a one-way ANOVA with four groups, at 95% power and an alpha level of .05. To account for the fact that our control group would be included in both sets of analyses, we doubled the size of the control group (thus the required sample size became *N* = 2155). We deliberately recruited a larger sample to account for the exclusion criteria detailed below.

Participants were recruited through online panel provider Respondi (respondi.com) between 28 January-1 February 2021, and quota sampled to reflect the UK population in terms of age and sex. Individuals who did not provide informed consent, reported already having received a COVID-19 vaccination, or did not live in the UK were screened from taking the survey. A total of 2,509 participant’s completed the survey. As per pre-registration, we excluded participants who failed an attention check (‘please select somewhat agree’; n = 290), or provided an age outside of the range of 18-100 years old (n = 2). This resulted in a final sample of 2,217 participants (47% males; *M*_age_ = 46.33, *SD* = 15.76; 88% White ethnicity; median highest education level: school education up to age 18).

### Materials and procedure

After providing consent, participants were immediately randomised by the Qualtrics randomisation tool to one of seven conditions, to which they were blinded (with twice as many participants randomised to control as to each of the other conditions). Participants in the different information conditions (but not control) were told they would be shown ‘a short piece of information about the COVID-19 vaccines’ and asked to answer questions about it, including some testing recall. After completing the questions regarding COVID-19 vaccines, all participants completed a series of demographic questions.

The six messages used in this study were modelled closely on publicly available communications available at the time of the study, including information pages from the Public Health England (2021b) website, and tweets from the UK NHS (NHS, 2021) and Public Health England (2021a) twitter accounts. We developed three types of message: No Caution, Medium Caution and High Caution. **No caution** messages highlighted the efficacy and safety of vaccines but made no mention of the need to continue protective behaviours once vaccinated. **Medium Caution** messages stated that while vaccines are broadly safe and effective, there is some uncertainty, and noted the need to maintain protective behaviours after vaccination. **High Caution** messages emphasized uncertainty around vaccine efficacy and noted the need to maintain protective behaviours after vaccination. For each type we created a **Long** version reflecting what might be found on a webpage, and a **Short** 2-3 sentence version formatted in the style of a social media post. All messages were presented with an identical image of vaccine vials. As an example, the Long of version of the Medium Caution message is shown in Figure 3. (Appendix 4 details the full text of all messages as presented to participants).

**Figure 3:**
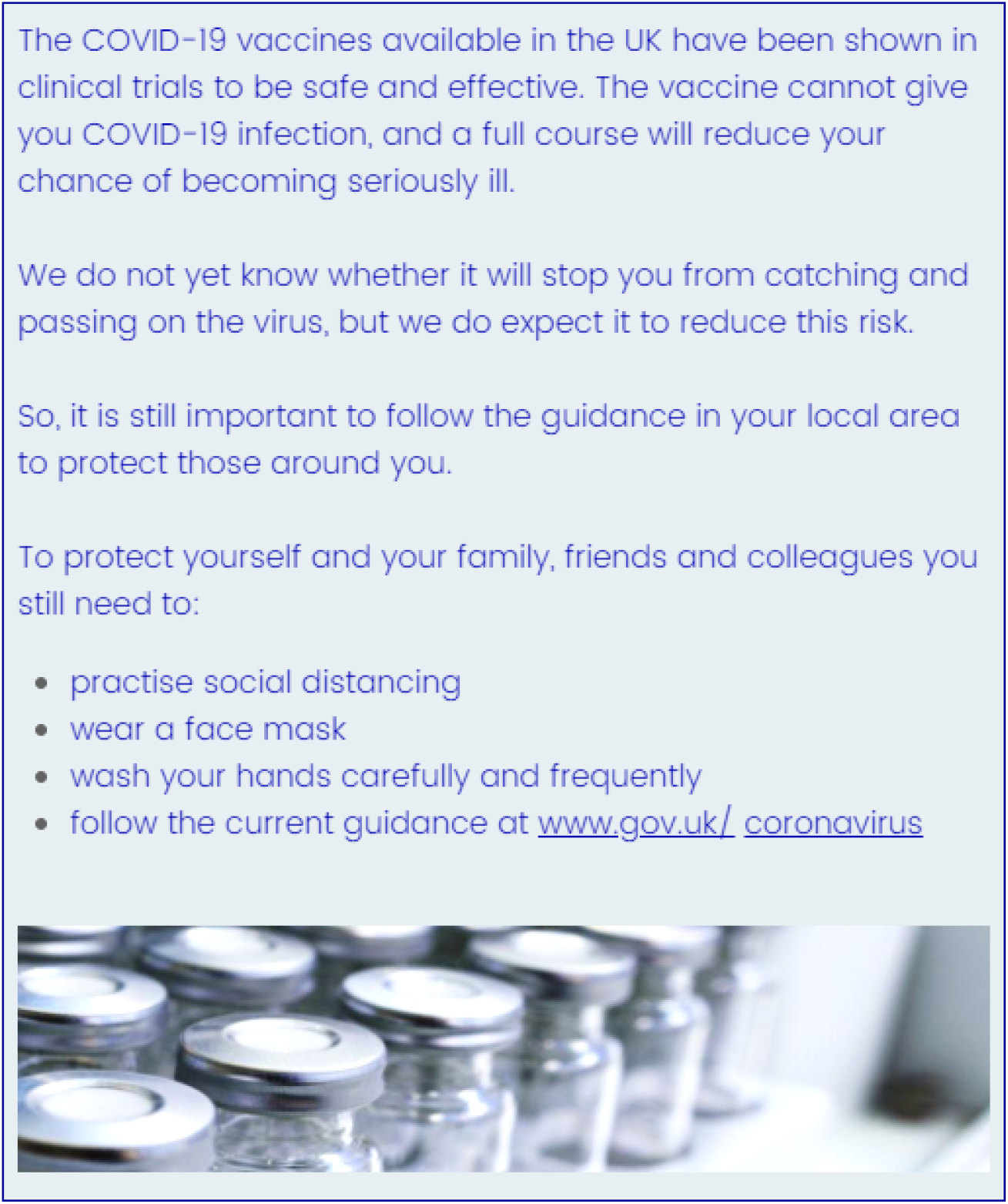
The Long Medium Caution message.

Participants then completed the following measures (in order presented; full item wording can be found in Appendix 5).

#### Primary Outcomes

Participants first completed the primary outcome measures. Intentions to engage in protective behaviour following vaccination were measured in two ways. First participants were asked to imagine they had been vaccinated and indicate how likely it is that they would engage in each of six behaviours once vaccinated (social distancing, frequent handwashing, wearing a facemask, avoiding social gatherings, working from home, avoiding public transport; 7pt response scale, *Very unlikely* to *Very Likely*). Responses were averaged to create a reliable index of protective behaviour intention (α = .86). Second, participants were asked if they would follow general rules post-vaccination, indicating their agreement with the statement: *I would still follow whatever coronavirus rules or restrictions were in place as strictly as I was before getting a vaccine* (7pt, *Strongly disagree* to *Strongly agree*). Willingness to receive a vaccine was measured using the Oxford COVID-19 Vaccine Hesitancy Scale (Freeman et al., 2021; α = .97; as in Study 1, scores were reversed such that higher values indicate greater willingness).

#### Secondary Outcomes

Participants also completed a second, binary measure of vaccination intention, responding *Yes* or *No* to the question: ‘*If you were offered a COVID-19 vaccine would you get vaccinated yourself?*’ Hypothetical worry over COVID-19 risk behaviours post-vaccination was measured with a set of items in which participants indicated their level of worry regarding seven different behaviours (e.g. *Shopping in a busy supermarket*) if they had already received the COVID-19 vaccine. Scores were averaged to create a single index (α = .95).

Participants who read a message (i.e. all groups other control) then completed several ‘message only’ measures relating to their impression of the information presented. Emotional response was captured using Marcus et al.’s (2017) nine-item scale in which participants indicate how the information made them feel (e.g. *Resentful*) on a slider with the labels *Not at all* (0) and *Extremely* (100) and the slider initially anchored at 50. Following Marcus et al., responses were averaged to create three subscales: anxiety (α = .92), aversion (α = .93), and enthusiasm (α = .85). These participants also completed measures of: how trustworthy (three items, α = .97), engaging (three items, α = .87), and believable they found the information; how much they felt they understood the information; the level of effort required to read the information; and the perceived quality of the evidence underlying the information (all identical to Study 1).

All participants then completed the adapted version of the perceived efficacy (α = .84) and public importance (example: *If I get the COVID-19 vaccine it will be:* 1 = *Really helpful for the community around me*, 5 = *Really unhelpful for the community around me*; α = .84) subscales of the Oxford COVID-19 Vaccine Beliefs Scale, the informed (α = .81) and certainty (α = .91) measures adapted from the Decisional Conflict Scale, and provided estimates of the efficacy of the ‘best available’ COVID-19 vaccines (percent of cases prevented; all measures identical to Study 1).

### Analysis

As per our preregistration, we compared outcome means between the long messages and control, and short messages and control separately using one-way ANOVAs (i.e. four groups in each separate analysis; hereafter differentiated by *Long* and *Short* subscripts in results).

## Results

As in Study 1, participants, on average, considered the messages to be relatively trustworthy (*M*s 5.46-5.71, *SD*s 1.28-1.47), engaging (*M*s 5.47-5.76, *SD*s 1.20-1.30) and believable (*M*s 5.41-5.66, *SD*s 1.29-1.46), with mean scores on these measures above the scale mid-point (range 1-7) across all message conditions (see Appendix 6 for comparisons between conditions).

### Primary outcomes

Comparing both Long and Short messages separately with control, condition assignment was not significantly associated with willingness to receive a vaccine (*F*_Long_(3, 1389) = 1.91, *p* = .13; *F*_Short_(3, 1396) = 2.10, *p* = .10, η2 = 0.00), intentions to follow general rules and restrictions (*F*_Long_(3, 1390) = 1.38, *p* = .25; *F*_Short_(3, 1396) = 0.66, *p* = .57), or to engage in specific behaviours following vaccination (*F*_Long_(3, 1390) = 2.28, *p* = .08; *F*_Short_(3, 1395) = 0.95, *p* = .42). See Figure 4.

**Figure 4:**
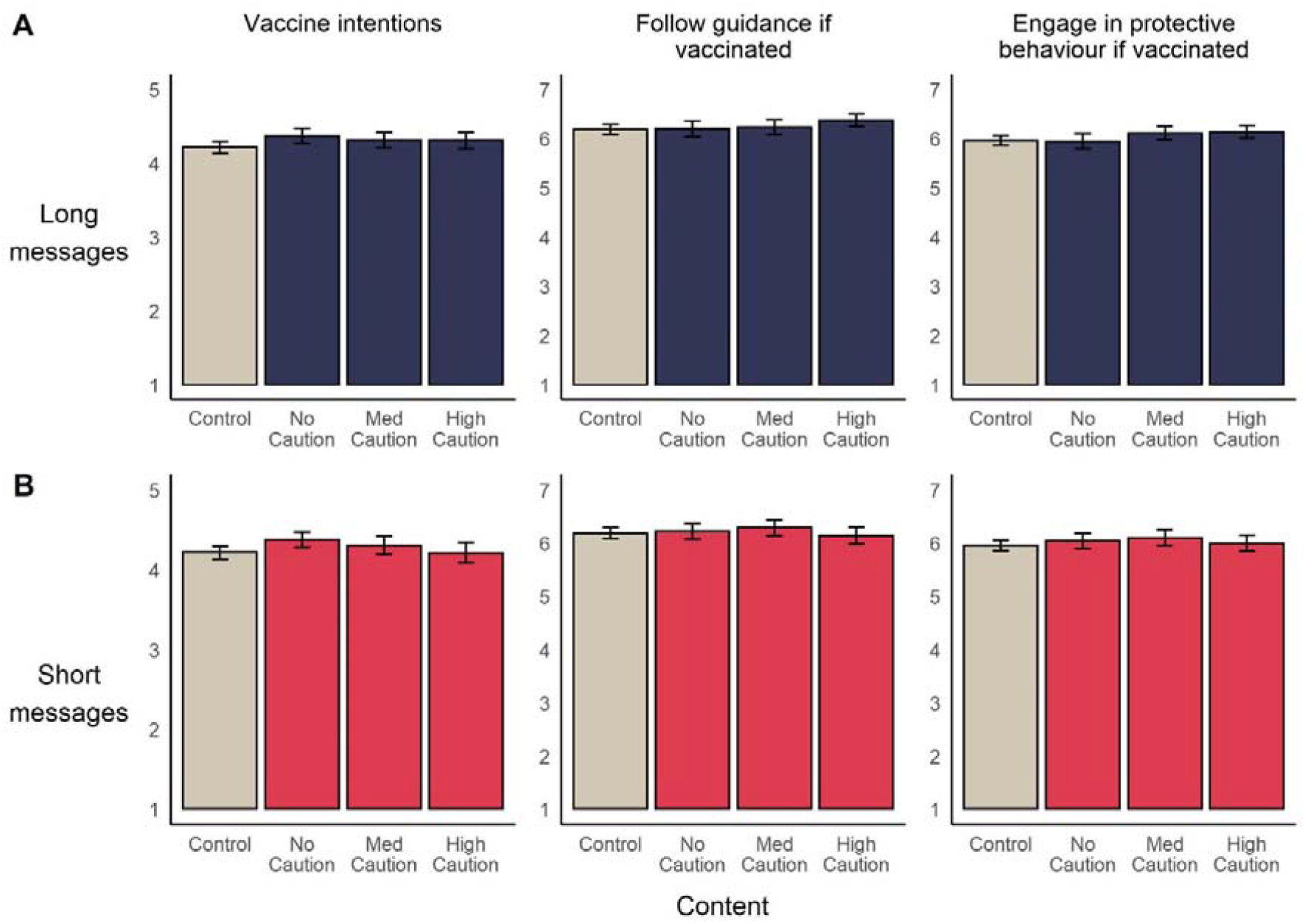
Mean (95%CI) levels of vaccine and behavioural intentions, across experimental conditions. In line with ANOVAs reported in main text, Long messages (A) and Short messages (B) are separately compared to control.

To examine extent to which the data supports a null effect model, we conducted Bayesian ANOVAs. Resulting Bayes factors indicated ‘substantial’ to ‘very strong’ evidence for null models in which message condition has no effect vaccine intentions, (*BF*_Long_ = 30.46; *BF*_*S*hort_ = 24.13), intentions to follow rules (*BF*_Long_ = 67.66; *BF*_*S*hort_ = 190.01) or to engage in protective behaviour post vaccination (*BF*_Long_ = 18.38; *BF*_*S*hort_ = 125.14).

### Secondary outcomes

As in Study 1, we included an alternative binary (yes/no) measure of vaccine intentions in the experiment. In the control group, 84.9% of participants indicated they would receive a vaccine if available. A chi-squared test of independence indicated that the proportion of ‘yes’ responses did not differ significantly across message conditions (*X*^2^(6)= 4.78, *p* = .57). Additional tests comparing only Long messages and the control group (*X*^2^(3)= 3.55, *p* = .31) and Short messages and the control group (*X*^2^(3)= 2.08, *p* = .56) were also non-significant.

We report no main effect of message condition on reported worry over risk behaviours if vaccinated (*F*_Long_(3, 1390) = 2.46, *p* = .06; *F*_Short_(3, 1396) = 0.72, *p* = .54), the perceived public importance of vaccines (*F*_Long_(3, 1390) = 1.87, *p* = .13; *F*_Short_(3, 1396) = 2.16, *p* = .09), or decision certainty (*F*_Long_(3, 1388) = 2.44, *p* = .06; *F*_Short_(3, 1394) = 1.87, *p* = .13).

Comparing between Long messages and the control group, we do find a significant effect of message on perceived efficacy of vaccines, *F*(3, 1389) = 7.76, *p* < .001, η^2^ = 0.02. Post hoc tests revealed that participants in the No Caution condition (*M* = 4.14, *SD* = 0.70) perceived COVID-19 vaccines to be more effective than participants in the control (*M* = 3.89, *SD* = 0.77; *p* < .001, *d* = .34), Medium Caution (*M* = 3.93, *SD* = 0.71; *p* < .01, *d* = .29), and High Caution (*M* = 3.92, *SD* = 0.73; *p* < .01, *d* = .31) conditions. There was a significant effect of condition on quantitative estimates of vaccine efficacy (i.e. estimated percent of cases prevented by vaccine), F(3, 1386) = 8.09; *p* < .001, η2 = 0.02, with the mean estimate in the No caution condition (*M* = 78.65%, *SD* = 20.76) significantly higher than that in the control (*M* = 70.99%, *SD* = 23.07; *p* < .001, *d* = .35) and Medium Caution (*M* = 73.75%, *SD* = 19.97, *p* = .04, *d* = .24) conditions. We also note a significant effect of condition on participants’ reported level of feeling informed regarding their vaccination decision, F(3, 1390) = 4.96, *p* < .01, η^2^ = 0.01, with average responses in the No Caution (*M* = 4.20, *SD* = 0.83, *p* = .02, *d* = .22) and Medium Caution (*M* = 4.22, *SD* = 0.81; *p* = .006, *d* = .25) conditions higher than that in control condition (*M* = 4.01, *SD* = 0.93), as shown in Figure 5.

**Figure 5:**
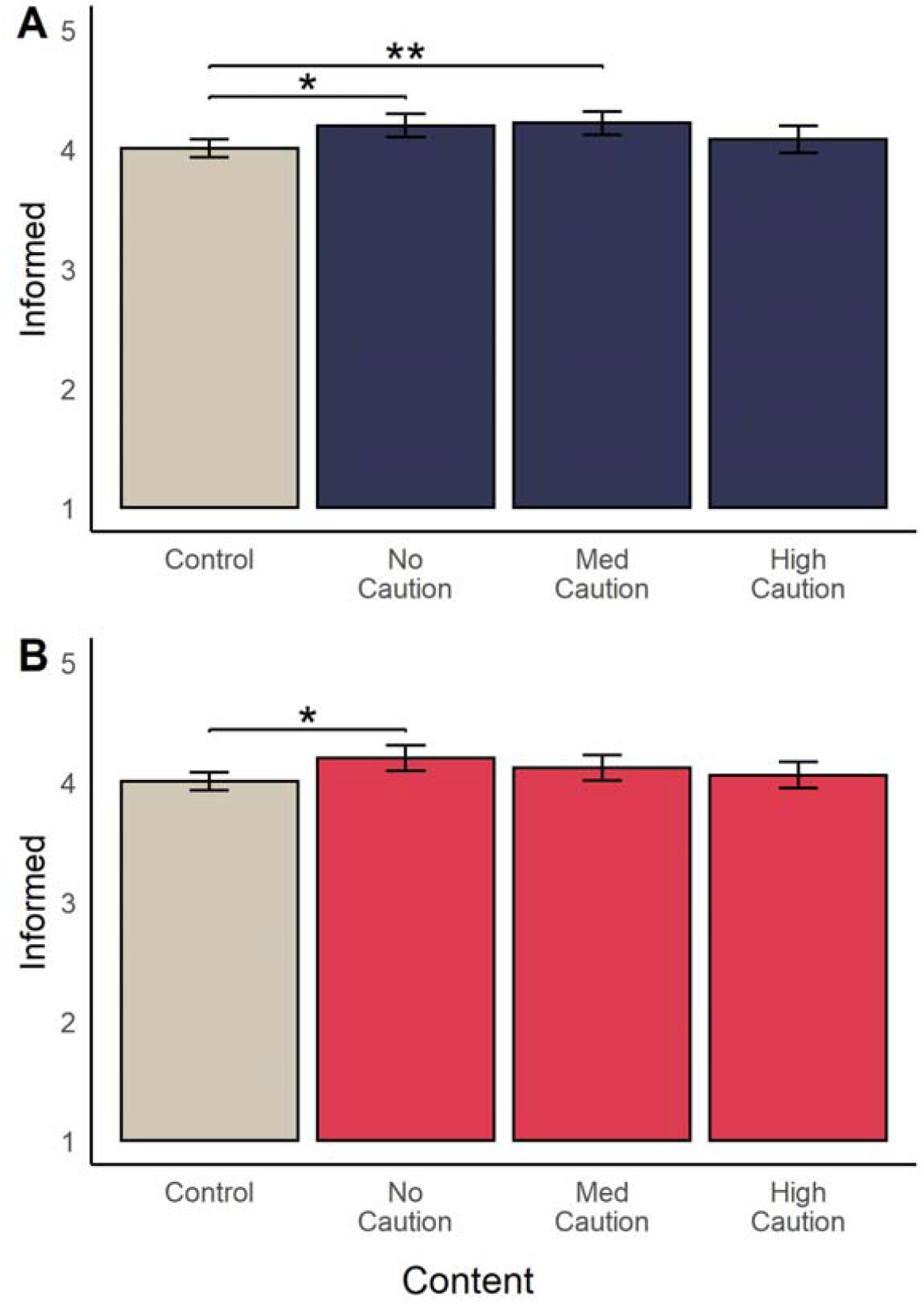
Mean (95%CI) reported level of feeling informed after reading (A) Long and (B) Short messages. Brackets and asterisks indicate significant pairwise differences between conditions. \**p* < .05, \*\**p* < .01.

Comparing Short messages and the control, we also report a significant effect of condition on perceived efficacy (*F*(3, 1394) = 4.86, *p* < .01, η2 = 0.01) and estimated efficacy (F(3, 1392) = 8.59, *p* < .001, η^2^ = 0.02). Post hoc tests indicated that perceived efficacy was significantly higher in the No Caution condition (*M* = 4.05, *SD* = 0.72) compared to the control (*M* = 3.89, *SD* = 0.77; *p* = .02, *d* = .21 and High Caution conditions (*M* = 3.86, *SD* = 0.83; *p* = .03, *d* = .23). For quantitative estimates of vaccine efficacy, we find that estimates in the No Caution condition (*M* = 76.10%, *SD* = 20.00) were higher than those in the control condition (*M* = 70.99%, *SD* = 23.07; *p* < .01, *d* = .24), as were estimates in the Medium Caution condition (*M* = 78.69%, *SD* = 20.95; *p* < .001, *d* = .35). The ANOVA test of the effect of condition on feeling informed was significant, *F*(3, 1396) = 3.09, *p* = .03, η^2^ = 0.01: participants’ in the Caution condition (*M* = 4.20, *SD* = 0.92) reported feeling more informed than those in the control condition (*M* = 4.01, *SD* = 0.93; *p* = .02, *d* = .21; Figure 4).

There were a number of measures relating to perceptions of the information shown which were not presented to participants in the control condition (because these participants did not see any message). For these outcomes, we conducted 2(length)x3(content) ANOVAs to examine the effect of both message length and content. Briefly, we find there was no significant effect of either factor, or interaction between them, on: perceived trustworthiness of the information; perceived quality evidence upon which the message was based; how believable the information was; how difficult the information was to understand. Longer messages were considered more engaging than short messages. Considering emotional responses to messages, No Caution messages elicited more enthusiasm and less aversion and anxiety compared to other conditions (full results reported in Appendix 6).

#### Study 2 discussion

As in Study 1, we find that intentions to vaccinate remain relatively static in the face of messages about vaccine efficacy and safety. We also find that the inclusion of uncertainty and reminders to maintain protective behaviours once vaccinated do not have a substantial effect on intentions to vaccinate, or on intentions to engage in protective behaviours if vaccinated. This will be of some comfort to public health communicators concerned that calls to continue social distancing and mask-wearing once vaccinated–another important public health message (Su et al., 2021)—might undermine perceptions of vaccine efficacy.

Considering our secondary outcomes, we did find that positive (i.e. No Caution) messages did increase the subjective perceived efficacy of the vaccines and quantitative estimates of vaccine efficacy, relative to the control group. Similarly, both Long and Short No caution messages (and the Long Medium caution message) increased participants’ sense of feeling informed regarding their vaccination decision compared to the control group. This is an interesting comparison with Study 1, where it was the provision of more informative messages that increased reported informedness. Here in Study 2 it is the *least* informative messages that increased the subjective impression of being informed. This raises the issue of self-report knowledge appraisals not being a good measure of decision quality when taken in isolation: a decision made easy through simplified or persuasive messaging, which might lead to high rating on decision confidence, is not necessarily a good decision.

## General discussion

The clear and transparent communication of COVID-19 vaccines’ risks and benefits has been suggested as an approach to increasing vaccine uptake among the general public (Cohen et al., 2020; Rzymski et al., 2021), a means of maintaining public trust in science (e.g., Nature, 2020; Parmet, 2005), and also as an ethical imperative. We investigated how communications about the efficacy and side effects of COVID-19 vaccines affect individuals’ vaccination intentions and related beliefs. Across two experiments we find that intention to receive a vaccine when offered is relatively high and that different messages—ranging from detailed tables of vaccine efficacy and side effects to a simple claim that vaccines are safe and effective—do not substantially increase intentions to receive a COVID-19 vaccine. Encouragingly, we also did not find any evidence that such communications lead to a decrease in vaccination intentions either, such as through ‘backfire effects’ (Wood & Porter, 2019). This was true even for messages in Study 2 which expressed high uncertainty about vaccine efficacy (a range of “41-98%”) and reminded the reader that protective behaviours would need to be maintained even once vaccinated.

We also examined how messages influenced participants’ perceptions of COVID-19 vaccine efficacy and safety, beliefs which are consistently associated with vaccine intentions (Freeman et al., 2021; C. Lin et al., 2021). These beliefs remained essentially unchanged in the face of detailed information from clinical trials. We do note that in Study 2, simple claims that vaccines are safe and effective (with or without reference to specific details; i.e. both short and long versions) did increase perceptions of efficacy relative to controls, but had no main effect on vaccine intentions. This gives some indirect evidence that increasing perceptions of vaccine efficacy may have a limited subsequent impact on intentions. However further research should investigate the causal effects of changes in perceived efficacy on intentions to confirm this.

Taken as whole, our main results align with the wider research on non-COVID-19 vaccine messaging, which indicates that simple written or visual purely informational messages have limited effects on vaccine hesitancy and uptake (European Centre for Disease Prevention, 2017; Penta & Baban, 2018). Our results also indicate that COVID-19 vaccine hesitancy is not simply due to “a deficit of the data needed to make an informed decision,” as suggested by Opel et al., (Opel et al., 2020). While the current studies do not provide a strong basis for public health authorities to communicate vaccine information with the aim of increasing uptake of the vaccine, we would note two important points: first our study occurred in the midst of such communications, and second, increasing uptake is not the only reason for actively disseminating vaccine evidence such as the results of clinical trials.

On the first point, public health bodies in the UK, where these studies were carried out, have been active in communicating information about COVID-19 vaccines as it becomes available, and such information has received intense media attention. This may have already contributed to the high levels of vaccination intentions. As such, we may be constrained by a celling effect in the current study. We did not measure attitudes prior to the experimental interventions, but it is possible that there was a greater effect of information on people who were opposed to or undecided regarding COVID-19 vaccination. Future research could investigate such moderation effects, incorporating prior beliefs into experimental models or using pre-post within subjects designs.

Regarding the second point, governments and responsible public health bodies have an ethical duty to inform the public of the risks and benefits of the vaccines they are asking the public to take (J. O’Neill, 2020). Thus informing the public, as much as persuading the public, should be considered an important endpoint. We did find some positive results in this regard; in Study 1 several of the more informative messages increased participants’ sense of feeling informed regarding the decision of whether or not to vaccinate. Such transparent communication may also serve to build trust over the longer term (Balog-Way & McComas, 2020; Larson & Heymann, 2010).

One notable omission from the various the framings included in this study was a focus on prosocial or collective benefits of vaccination. At the time of data collection, there was no peer-reviewed research indicating that COVID-19 vaccines could prevent transmission of the virus. As such, we did not include any messages detailing how the choice to vaccinate might protect others. Previous research on hesitancy towards other vaccines (Betsch et al., 2017; Betsch & Böhm, 2018) and preliminary studies on COVID-19 vaccine intentions (Motta et al., 2021; Williams et al., 2020) suggest that this framing may be effective in increasing vaccine acceptance. As more evidence of the effect of vaccination on transmission becomes available, researchers should investigate the how communicating such evidence impacts vaccine attitudes and intentions. This may be especially relevant for increasing vaccination rates in younger age groups who are at much lower risk of death or hospitalisation from COVID-19 and perceive less threat from the virus (Karlsson et al., 2021; Troiano & Nardi, 2021). Another factor we did not examine in the current studies was the impact of message source. It is possible that attributing the information to a source considered trustworthy by the UK public (e.g. the NHS) may have increased the impact of information.

Beyond broad messaging approaches, those seeking to increase vaccine uptake have a range of tools at their disposal. Interventions such as messaging targeted specifically to hesitant groups, working with trusted community leaders, training healthcare worker for face-to-face conversations with vaccine hesitant individuals, and reducing financial or logistical barriers (e.g. access to vaccination sites) have all been shown to increase vaccine uptake in some contexts (European Centre for Disease Prevention, 2017; Paterson et al., 2016; WHO, 2014). A comprehensive approach to addressing COVID-19 vaccine hesitancy should draw on all available tools, and not rely solely on communicating vaccine information via messages like those examined in the current studies.

We must note some limitations to the current studies. Both used online panel samples, which may not be reflective of the wider UK population. However, we would note that our samples were matched to the UK population in terms of age and gender, and the proportion of participants willing to receive a vaccine was in line with weighted population estimates reported in a contemporaneous YouGov survey (88.8% in February 2021; Ansell et al., 2021). Readers should also be mindful that results from our UK studies may not generalise to other countries, as many local contextual factors can influence vaccine hesitancy (WHO, 2014). For example, the development of the Oxford AstraZeneca vaccine has been hailed by the media as a ‘British’ success story (e.g., Nuki, 2020) and this may lead to pro-vaccine attitudes being linked with national identity in the UK.

In conclusion, we find that messages outlining the known risks and benefits of COVID-19 vaccines (with varying levels of detail) have little impact on vaccination intentions or beliefs about COVID-19 vaccines in UK online samples but some messages increase reported understanding of the risk and benefits. We also found no evidence that emphasising the need for continuing protective behaviours post-vaccination undermined vaccination intentions or perceptions of efficacy. These results indicate that there are no substantial effects of transparent communication on reducing vaccination intentions. However, they also highlight the need for those who wish to *increase* vaccination intentions or protective behaviours post vaccination to draw on a range of strategies—not just transparent communication of risk and benefits.

## Data Availability

Data and analysis code for these studies are available at: https://osf.io/hsugy/?view_only=16fbf1f5737a43db8f9d20a7b2d3db4b

## Acknowledgements

We would like to thank Lisa-Maria Tanase for assistance in developing and proofing survey materials.

## Footnotes

1 The six Decisional Conflict Scale (DCS) items were selected as the most relevant to the COVID-19 vaccine decision and we had intended treat these as a single scale (α = .90). However parallel and exploratory factor analyses revealed two factors differentially impacted by the experimental intervention. This lead to the creation of the two ad hoc two scales reported here (a deviation from our pre-registration). With regards to the original DCS factor structure, the three certainty items come from the Uncertainty subscale, whereas the informed items in the current study comprise two items from the Informed subscale (knowing the risks and benefits) and one from the Effective Decision subscale (‘I would feel I had made an informed choice’).

2 As estimates of severe side effect frequency displayed notable skew (> 4 across all conditions), we also conducted non-parametric tests. We note this is a deviation from our pre-registered analyses. A Kruskal-Wallis test indicated a significant effect of condition on estimates, *H*(4) = 11.11, *p* = .02. Post-hoc comparisons using Dunn’s Test indicated that the median estimate in the Approval condition (Median = 20) was significantly lower than the median estimate in the Factbox condition (Median = 50; *p* = .02), but no message condition differed significantly from the control group.

