## Supplementary information for "Effect of information about COVID-19 vaccine effectiveness and side effects on behavioural intentions: two online experiments"

### Supplementary material

### Appendix 1: Study 1 messages

This section details the full text of the messages presented to participants in Study 1.

***Factbox Condition***

---

**Information from the clinical trial of the new COVID-19 vaccine**

Several new vaccines have been developed to protect people against COVID-19, which is the disease caused by the SARS-Cov2 coronavirus. These vaccines have been developed and tested throughout 2020 and are now becoming available to the public.

Here we show the information on the benefits (reduction in COVID-19 cases) and harms (increased side effects and adverse reactions) for one of the new vaccines now available and licensed.

The numbers come from the study, in which volunteers were given either two doses of the vaccine or two dummy injections (placebo). We have split the information into two age groups (18-64 and over 65). Under 18s were not involved in the vaccine testing.

So far volunteers have been followed for a maximum of 8 months. Monitoring and reporting of serious adverse reactions will continue as the vaccine is rolled out to the public.

The results from the study (as of 7th December 2020) are:

**For 18-64 year olds:**

| POTENTIAL BENEFITS<br>From 2 weeks after 2nd dose | | | |
|---|---|---|---|
| | **Dummy injection (10,521 people)** | **Vaccine injection (10,551 people)** | **What difference did the vaccine make?** |
| Number who developed symptoms confirmed to be COVID-19 | 156<br>(1.5%) | 7<br>(less than 0.1%) | 149 fewer cases<br>(95.5% reduction in COVID-19 cases) |

| POTENTIAL HARMS*<br>(usually lasting 2-3 days) | | |
|---|---|---|
| Number who reported: | **Dummy injection (10,315 people)** | **Vaccine injection (10,357 people)** |
| Pain at the injection site<br>(some also reported redness and swelling) | 1,942<br>(18.8%) | 9,335<br>(90.1%) |
| Swollen/sore armpit glands | 444<br>(4.3%) | 1,654<br>(16%) |
| Fever | 38<br>(0.4%) | 1,806<br>(17.4%) |
| Headache<br>(a similar number reported other 'flu-like symptoms such as fatigue, aching joints, chills) | 2,617<br>(25.4%) | 6,500<br>(62.8%) |
| Nausea/Vomiting | 754<br>(7.3%) | 2,209<br>(21.3%) |

For 65+ year olds:

| POTENTIAL BENEFITS<br>From 2 weeks after 2nd dose | | | |
|---|---|---|---|
| | **Dummy injection (3,552 people)** | **Vaccine injection (3,583 people)** | **What difference did the vaccine make?** |
| Number who developed symptoms confirmed to be COVID-19 | 29<br>(0.8%) | 4<br>(0.1%) | 25 fewer cases of COVID-19<br>(86.2% reduction) |

| POTENTIAL HARMS*<br>(usually lasting 2-3 days) | | |
|---|---|---|
| Number who reported: | **Dummy injection (3,552 people)** | **Vaccine injection (3,583 people)** |
| Pain at the injection site<br>(some also reported swelling and redness) | 421<br>(11.9%) | 2,990<br>(83.4%) |
| Swollen/sore armpit glands | 90<br>(2.5%) | 302<br>(8.4%) |
| Fever | 5<br>(0.1%) | 366<br>(10.2%) |
| Headache<br>(a similar number reported other 'flu-like symptoms such as fatigue, aching joints, chills) | 635<br>(17.9%) | 1,665<br>(46.4%) |
| Nausea/Vomiting | 129<br>(3.6%) | 425<br>(11.8%) |

**More serious side effects for all age groups**

As well as these short-lasting side effects, the trial recorded any serious medical events that people of all ages in the trial experienced in 28 days after they got the injections. The events where people in the vaccine group suffered more than those in the dummy injection group were:

| | **Dummy injection (13,906 people)** | **Vaccine injection (13,911 people)** |
|---|---|---|
| Heart attack | 3 | 5 |
| Inflammation of gallbladder | 0 | 3 |
| Kidney stones | 0 | 3 |
| Bell's palsy | 1 | 3 |
| We also can't be sure whether the vaccine did or did not contribute to the following: | | |
| Rheumatoid arthritis | 0 | 1 |
| Swelling or shortness of breath when exercising | 0 | 1 |
| Autonomic dysfunction eg dizziness when standing, sweating too much or too little, heart rate changes | 0 | 1 |

* Potential harms data is from the 2nd dose injection. Data from the 1st dose injection shows very slightly lower numbers of each reaction.

**Information from the clinical trial of the new COVID-19 vaccine**

Several new vaccines have been developed to protect people against COVID-19, which is the disease caused by the SARS-Cov2 coronavirus. These vaccines have been developed and tested throughout 2020 and are now becoming available to the public.

The vaccines are given as two injections, usually into the muscle of the upper arm.

Here we present information about one of the vaccines now approved by the regulators and available to the public:

**What benefits of the vaccine have been shown in studies?**

A very large clinical trial showed that the vaccine was effective at preventing COVID 19 in people from 18 years of age. The trial involved around 30,000 people in total. Half received the vaccine and half were given dummy injections. People did not know whether they received the vaccine or the dummy injections.

Efficacy was calculated in around 28,000 people from 18 to 94 years of age who had no sign of previous infection. The trial showed a 94.1% reduction in the number of symptomatic COVID-19 cases in the people who received the vaccine (11 out of 14,134 vaccinated people got COVID-19 with symptoms) compared with people who received dummy injections (185 out of 14,073 people who received dummy injections got COVID-19 with symptoms). This means that the vaccine demonstrated a 94.1% efficacy in the trial.

The trial also showed 90.9% efficacy in participants at risk of severe COVID-19, including those with chronic lung disease, heart disease, obesity, liver disease, diabetes or HIV infection.

**Can the vaccine reduce transmission of the virus from one person to another?**

The impact of vaccination on the spread of the SARS-CoV-2 virus in the community is not yet known. It is not yet known how much vaccinated people may still be able to carry and spread the virus.

**How long does protection from the vaccine last?**

It is not currently known how long protection given by COVID-19 Vaccine Moderna lasts. The people vaccinated in the clinical trial will continue to be followed for 2 years to gather more information on the duration of protection.

**Can children be vaccinated with COVID-19 Vaccine Moderna?**

The COVID-19 vaccine is not currently recommended for use in children. The regulators have agreed with the company on a plan to conduct trials involving children at a later stage.

**Can people with allergies be vaccinated?**

People who already know they have an allergy to one of the components of the vaccine listed on the package leaflet should not receive the vaccine.

Allergic reactions (hypersensitivity) have been seen in people receiving the vaccine. A very small number of cases of anaphylaxis (severe allergic reaction) have occurred. Therefore, as for all vaccines, the COVID-19 vaccine should be given under close medical supervision, with the appropriate medical treatment available in case of allergic reactions. People who have a severe allergic reaction when they are given the first dose of the vaccine should not receive the second dose.

**How well does the vaccine work for people of different ethnicities and genders?**

The clinical trial included people of different ethnicities and genders. The high efficacy was maintained across genders and racial and ethnic groups.

**What are the risks associated with the vaccine?**

The most common side effects with the COVID-19 vaccine in the trial were usually mild or moderate and got better within a few days after vaccination. They included pain and swelling at the injection site, tiredness, chills, fever, swollen or tender lymph nodes under the arm, headache, muscle and joint pain, nausea and vomiting. They affected more than 1 in 10 people.

Redness, hives and rash at the injection site and rash occurred in less than 1 in 10 people. Itching at the injection site occurred in less than 1 in 100 people. Swelling of the face, which may affect people who had facial cosmetic injections in the past, and weakness in muscles on one side of face (acute peripheral facial paralysis or palsy) occurred rarely, in less than 1 in 1000 people.

Allergic reactions have occurred in people receiving the vaccine, including a very small number of cases of severe allergic reactions (anaphylaxis). As for all vaccines, the COVID-19 vaccine should be given under close supervision with appropriate medical treatment available.

**Information from the clinical trial of the new COVID-19 vaccine**

Several new vaccines have been developed to protect people against COVID-19, which is the disease caused by the SARS-Cov2 coronavirus. These vaccines have been developed and tested throughout 2020 and are now becoming available to the public.

COVID-19 vaccines can only be approved and used if they comply with all the requirements of **quality**, **safety** and **efficacy** set out in legislation. In view of the pandemic, regulatory agencies are diverting **resources** to speed up processes and reduce **timelines** for the evaluation and authorisation of COVID-19 vaccines.

Pharmaceutical legislation ensures that vaccines are only approved after scientific evaluation has demonstrated that their overall benefits outweigh their risks.

A vaccine's benefits in protecting people against COVID-19 must be far greater than any **side effect** or potential risks.

Scientific **experts** evaluating medicines do not have any financial or other interests that could affect their impartiality. A high level of transparency, which opens EMA’s scientific evaluation work to public scrutiny, safeguards the **independence** of EMA's scientific evaluations.

**Scientific evaluation and approval processes**
To gain approval for a vaccine in the EU, the vaccine developer submits the results of all testing / investigations to the medicines regulatory authorities in Europe. This is part of a marketing authorisation application.

Like all medicines, COVID-19 vaccines are first tested in the **laboratory** (e.g. studies on their pharmaceutical quality and studies to check first the effects in laboratory tests and animals).
Then vaccines are tested in human **volunteers** in studies called clinical trials. These tests help confirm how the vaccines work and, importantly, to evaluate their **safety** and protective **efficacy**.

**Standard vaccine development**
Standard vaccine development is a long process and studies are done in sequential steps.

Companies first make **small batches** and do small scale studies to characterise and optimise the production process. They perform studies to determinate a suitable formulation that can keep vaccine components **stable** to the end of its shelf life.

Then the company decides whether to continue development and scale up production. To assure that the vaccine meets its intended quality profile and complies with regulatory standards, the company develops a suitable and effective **quality control** strategy.

Studies on pharmaceutical quality look at the individual vaccine components, the final formulation to be used and at the whole **manufacturing process** in detail.

The vaccine developer conducts more studies in laboratory models, using **in vitro** studies or animal models (**in vivo** studies), to show how the vaccine triggers an immune response and works to prevent infection.

Finally, the vaccine developer studies the vaccine in three phases of **clinical trials**, with larger numbers of volunteers in each phase.

**Fast-track vaccine development in a public health emergency**

Vaccine development for COVID-19 vaccines is being fast-tracked globally. Development is compressed in time, applying the extensive knowledge on vaccine production gained with existing vaccines.

Early scientific advice from regulators helps speed up development. EMA offers informal consultation with its COVID-19 Task Force (ETF) and rapid scientific advice. COVID-19 vaccine developers can receive prompt guidance and direction on the best methods and study designs to generate robust **data**.

Advising companies on regulatory requirements helps ensure that standards of **quality**, **safety** and **efficacy** are embedded early in the process and are not compromised by fast-track development.

Regulators can also use a **rolling review** procedure for promising medicines for COVID-19. This allows EMA to begin assessing data as they become available during the development process, to expedite the subsequent formal marketing authorisation application assessment even further.

After approval, a larger number of people will receive the vaccine. Certain rare or very rare **side effects** may only emerge when millions of people are vaccinated. Law requires that the safety of vaccines is monitored while they are in use.

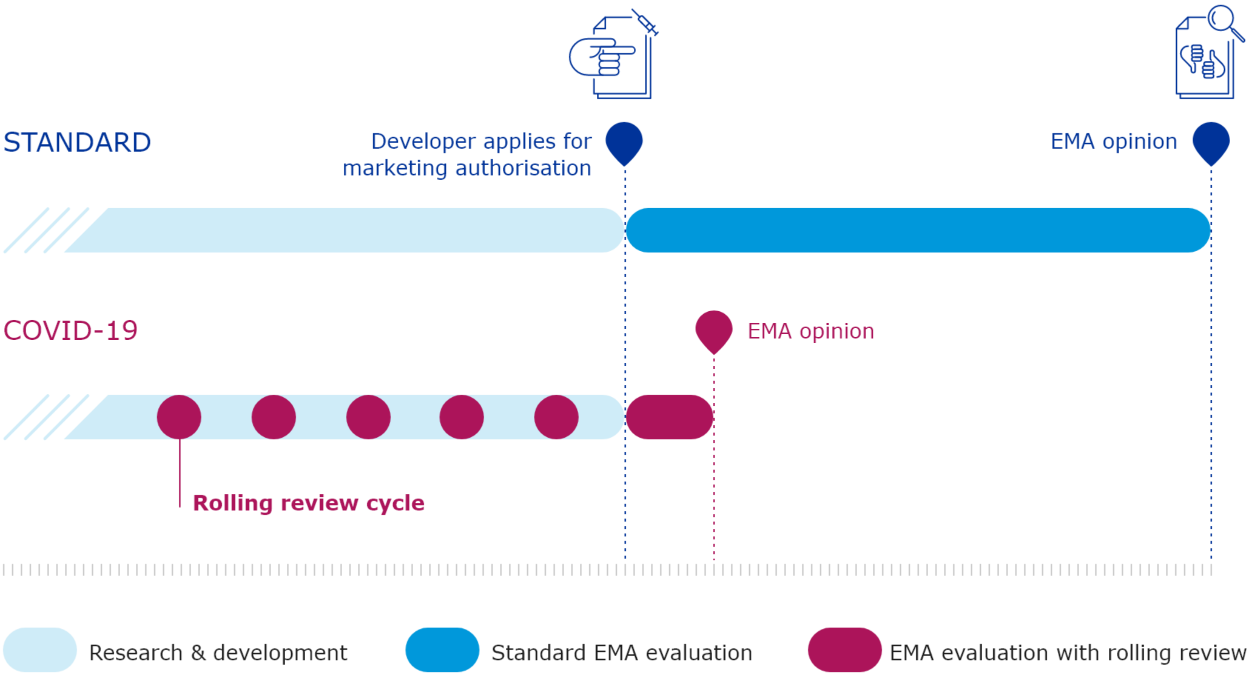
STANDARD
Developer applies for marketing authorisation
EMA opinion
COVID-19
EMA opinion
Rolling review cycle
Research & development
Standard EMA evaluation
EMA evaluation with rolling review

*Mechanism condition*

---

**Information on the new COVID-19 vaccine**

Several new vaccines have been developed to protect people against COVID-19, which is the disease caused by the SARS-Cov2 coronavirus. These vaccines have been developed and tested throughout 2020 and are now becoming available to the public.

Some of these vaccines are of a new type. Here we explain how they work:

Vaccines are considered one of the greatest developments of modern medicine, helping to nearly wipe out many infectious diseases. But creating and developing vaccines involves a long and complex process that remains a combination of art and science.

Our natural immune system is a network of cells, tissues and organs that work together to help fight off infection from harmful bacteria or viruses. When a disease-causing agent, such as virus or bacteria, invades your body, your **immune system** recognises it as harmful and will trigger a response to destroy it.

One of the ways your immune system fights off infection is by creating large proteins known as **antibodies**. These antibodies act as scouts, hunting down the infectious agent, and marking it for destruction by the immune system. Each antibody is specific to the bacteria or virus that it has detected and will trigger a specific immune response. These specific antibodies will remain in the immune system after the infection has gone. This means that if the same disease is encountered again, your immune system has a 'memory' of the disease and is ready to quickly destroy it before you get sick and any symptoms can develop.
Sometimes, however, the immune system doesn't always win this initial battle against the harmful bacteria or virus and you can become very ill or – in extreme cases – die.

The key to building this immunity is that the portion of the disease-causing agent called the **antigen** trains the immune system to recognise and respond to the agent. The antigen used to train the immune system against the virus that causes COVID-19 is a bit of its harmless outside coating called the "spike protein". When our immune systems recognize that the "spike protein" doesn't belong in the body, they begin building an immune response and making antibodies.

Many standard vaccines work by injecting a dead or weakened form of the disease-causing agent into the body in preparations that are designed not to make you sick but rather to build immunity by having the crucial antigen on the outside but not of the dangerous disease material on the inside.

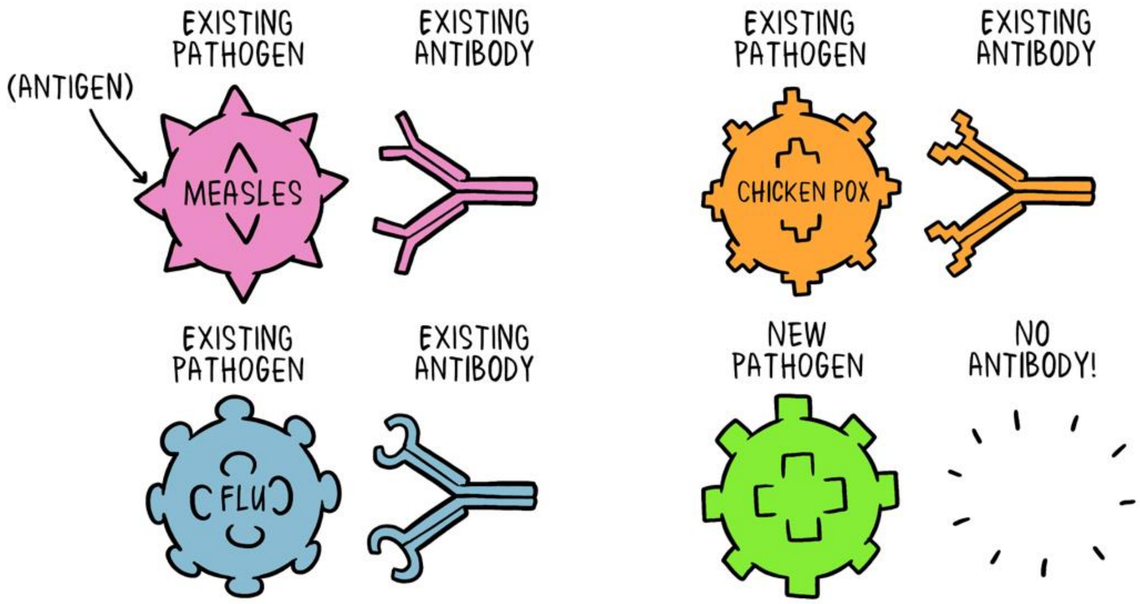


When a new pathogen or disease enters our body, it introduces a new antigen. For every new antigen, our body needs to build a specific antibody that can grab onto the antigen and defeat the pathogen.

However, instead of growing up harmless or weakened viruses that have the specific coronavirus “spike protein” in their outside coating and then injecting that into our bodies, the new COVID-19 vaccines instead contain a molecule called “messenger RNA” (or ‘mRNA’ for short). This is a short message, made in a laboratory, that, when injected into the body, tells some of our own immune cells to make the harmless “spike protein” instead. Once the “spike protein” is made, the cell breaks down the instructions and gets rid of them, just as it normally would. The person’s immune system will then recognise this “spike protein” as foreign and produce antibodies to attack it. If, later on, the person comes into contact with SARS-CoV-2 virus, their immune system will recognise the “spike protein” on its coating and be ready to defend the body against it.

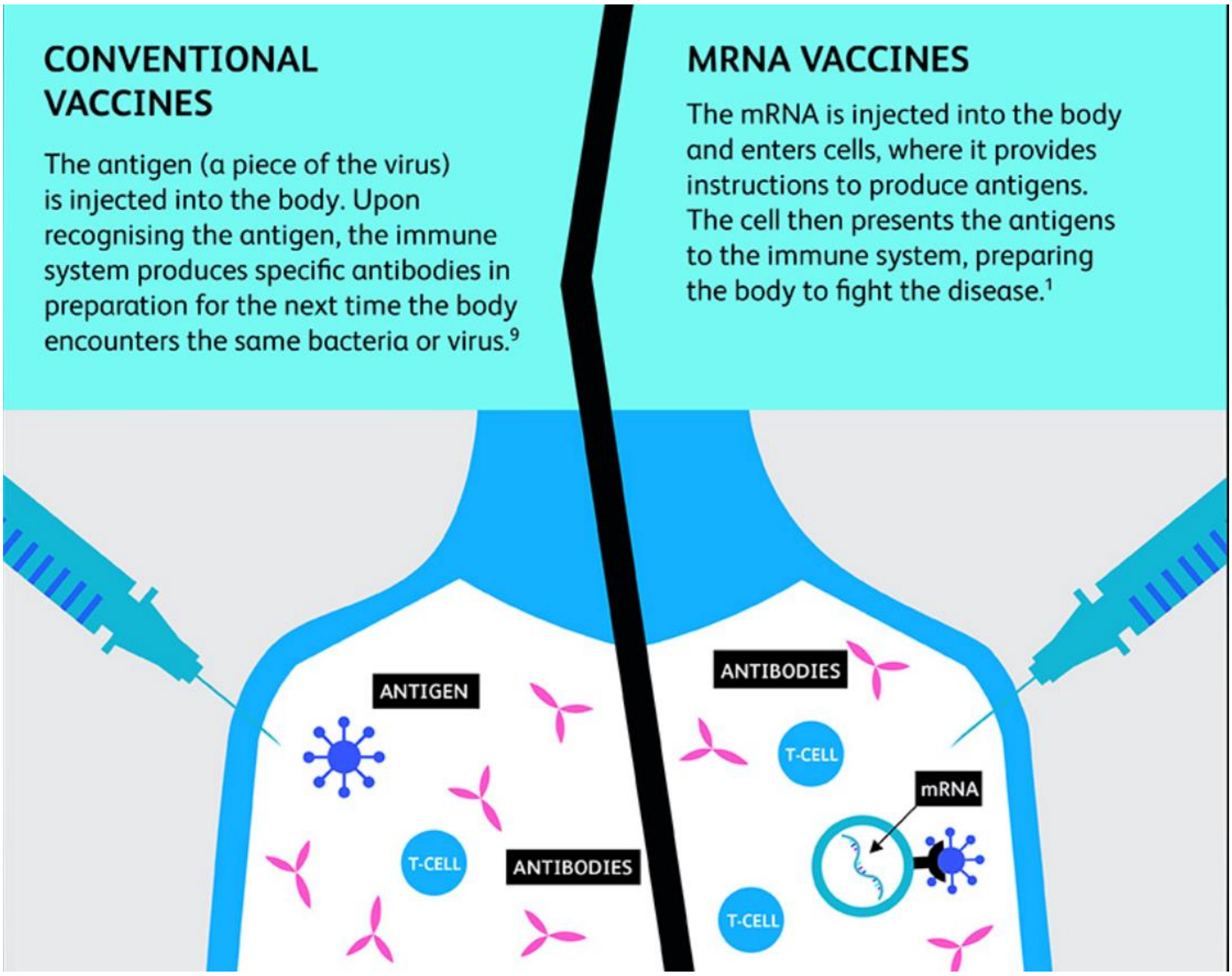


The mRNA from the vaccine does not stay in the body but is broken down shortly after vaccination and it is important to note that the mRNA strand never enters the cell's nucleus or affects genetic material. The vaccine does not contain the virus itself and cannot cause COVID-19.

Although the mRNA vaccines for COVID-19 are the first to be authorised, researchers have been studying them for decades. Early stage clinical trials using mRNA vaccines have been carried out for influenza, Zika, rabies, and cytomegalovirus (CMV). Because the mRNA material can be made in a laboratory, it avoids a lot of the slow and difficult process of 'growing up' and then deactivating dangerous viruses that are used in traditional vaccine development, making mRNA vaccines fast to design, produce and modify as new variants arise.

**Appendix 2: Study 1 survey items.**

Table S2.1
Survey items

| Scale | Label | Responses |
|---|---|---|
| Trustworthiness | Please tell us to what extent you think the information you just read was... - Accurate | Not at all (1) - Very much (7) |
| | - Reliable | Not at all (1) - Very much (7) |
| | - Trustworthy | Not at all (1) - Very much (7) |
| Quality of evidence | How high or low do you think the quality of the evidence underlying the information you just read is? | Low quality of evidence (1) - High quality of evidence (7) |
| Engagement | Still thinking about the information you just read, please answer the questions below. - In your opinion, would other people want to read this? | Not at all (1) - Very much (7) |
| | - Are you interested in this information? | Not at all (1) - Very much (7) |
| | - Do you like how this information is presented? | Not at all (1) - Very much (7) |
| Understanding | How completely do you feel you understood the information we just showed you? | Didn't understand it at all (1) - Understood it completely (7) |
| Effort | How much effort do you feel you had to put into understanding the information we just showed you? | None (1) - A lot (7) |
| Believability | How much did you believe the information you just read? | Didn't believe it at all (1) - Believed it completely (7) |
| Informed | If you were given the option of receiving a COVID-19 vaccine or not today how would you feel about the decision? - I know the benefits of getting vaccinated against COVID-19 | Strongly disagree (1) - Strongly agree (5) |
| | - I know the risks and side effects of getting vaccinated against COVID-19 | Strongly disagree (1) - Strongly agree (5) |
| | - I would feel I had made an informed choice | Strongly disagree (1) - Strongly agree (5) |

| | | |
|---|---|---|
| Certainty | - This decision is easy for me to make. | Strongly disagree (1) - Strongly agree (5) |
| | - I feel sure about what to choose. | Strongly disagree (1) - Strongly agree (5) |
| | - I am clear about the best choice for me | Strongly disagree (1) - Strongly agree (5) |
| Vaccination intention | Would you take a COVID-19 vaccine (approved for use in the UK) if offered? | Definitely (1)<br>Probably (2)<br>I may or I may not (3)<br>Probably not (4)<br>Definitely not (5) |
| | Now that there is a COVID-19 vaccine available in the UK: | I will want to get it as soon as possible (1)<br>I will take it when offered (2)<br>I'm not sure what I will do (3)<br>I will put off (delay) getting it (4)<br>I will refuse to get it (5) |
| | I would describe my attitude towards receiving a COVID-19 vaccine as: | Very keen (1)<br>Pretty positive (2)<br>Neutral (3)<br>Quite uneasy (4)<br>Against it (5) |
| | If a COVID-19 vaccine was available at my local pharmacy, I would: | Get it as soon as possible (1)<br>Get it when I have time (2)<br>Delay getting it (3)<br>Avoid getting it for as long as possible (4)<br>Never get it (5) |
| | If my family or friends were thinking of getting a COVID-19 vaccination, I would: | Strongly encourage them (1)<br>Encourage them (2)<br>Not say anything to them about it (3)<br>Ask them to delay getting the vaccination (4)<br>Suggest that they do not get the vaccination (5) |
| | I would describe myself as: | Eager to get a COVID-19 vaccine (1)<br>Willing to get the COVID-19 vaccine (2) |

| | | |
|---|---|---|
| | | Not bothered about getting the COVID-19 vaccine (3)<br>Unwilling to get the COVID-19 vaccine (4)<br>Anti-vaccination for COVID-19 (5) |
| | Taking a COVID-19 vaccination is: | Really important (1)<br>Important (2)<br>Neither important nor unimportant (3)<br>Unimportant (4)<br>Really unimportant (5) |
| Vaccination intention (binary measure) | If you were offered a COVID-19 vaccine: - Would you get vaccinated yourself? | Yes (1)<br>No (2) |
| Efficacy | Do you think you will be infected with COVID-19 over the next 12 months? (O) | Definitely (1)<br>Probably (2)<br>Possibly (3)<br>Probably not (4)<br>Definitely not (5) |
| | The COVID-19 vaccine is likely to: | Work for almost everyone (1)<br>Work for most people (2)<br>I am unsure how many people it will work for (3)<br>Not work for most people (4)<br>Not work for anyone (5) |
| | The COVID-19 vaccine is likely to: | Definitely work for me (1)<br>Probably work for me (2)<br>May or may not work for me (3)<br>Probably not work for me (4)<br>Definitely not work for me (5) |
| | The COVID-19 vaccine is effective in preventing COVID-19 (A) | Strongly agree (1)<br>Somewhat agree (2)<br>Neither agree nor disagree (3)<br>Somewhat disagree (4)<br>Strongly disagree (5) |

| | | |
|---|---|---|
| | People who are vaccinated against COVID-19 are less likely to get sick from the virus (A) | Strongly agree (1)<br>Somewhat agree (2)<br>Neither agree nor disagree (3)<br>Somewhat disagree (4)<br>Strongly disagree (5) |
| Concern over speed | The speed of developing and testing the vaccine means it will be: | Really good (1)<br>Good (2)<br>Will not affect how good or bad it is (3)<br>Bad (4)<br>Really bad (5) |
| | The speed of developing and testing the vaccine means it will be: | Really safe (1)<br>Safe (2)<br>It will not affect how safe it is (3)<br>Unsafe (4)<br>Really unsafe (5) |
| | Getting the vaccine is a sign of: (O) | Great personal strength (1)<br>Personal strength (2)<br>Not a sign of personal strength or weakness (3)<br>Personal weakness (4)<br>Great personal weakness (5) |
| | I have concerns about the speed at which the COVID-19 vaccine was developed and approved.*(A) | Strongly agree (1)<br>Somewhat agree (2)<br>Neither agree nor disagree (3)<br>Somewhat disagree (4)<br>Strongly disagree (5) |
| | COVID-19 vaccines have been approved too quickly *(A) | Strongly agree (1)<br>Somewhat agree (2)<br>Neither agree nor disagree (3)<br>Somewhat disagree (4)<br>Strongly disagree (5) |
| Concern over side effects | I expect that receiving the vaccine will be: | Hardly noticeable (1)<br>A little unpleasant (2)<br>Moderately unpleasant (3)<br>Painful (4)<br>Extremely painful (5) |
| | The side effects for people of getting the COVID-19 vaccine will be: | None (1)<br>Mild (2)<br>Moderate (3) |

| | | |
|---|---|---|
| | | Significant (4)<br>Life-threatening (5) |
| | Taking a new COVID-19 vaccine will make me feel like a guinea pig. | Do not agree (1)<br>Agree a little (2)<br>Agree moderately (3)<br>Agree a lot (4)<br>Completely agree (5) |
| | I am concerned about the potential side-effects of the vaccine.* (A) | Strongly agree (1)<br>Somewhat agree (2)<br>Neither agree nor disagree (3)<br>Somewhat disagree (4)<br>Strongly disagree (5) |
| Estimated mild to moderate side effect frequency | Mild to moderate side effects of vaccination include: pain and swelling at the injection site, tiredness, chills, fever, swollen or tender lymph nodes under the arm, headache, muscle and joint pain, nausea and vomiting.<br><br>In your own opinion, if 10,000 people were given a COVID-19 vaccine, how many of them do you think would experience the mild to moderate side effects described above?<br><br>Please enter your estimate in the box below. | [Numeric text entry; 0-10,000] |
| Estimated severe side effect frequency | In your own opinion, if 10,000 people were given a COVID-19 vaccine, how many of them do you think would experience severe side effects requiring medical attention?<br><br>Please enter your estimate in the box below. | [Numeric text entry; 0-10,000] |
| Estimated efficacy | By how much do you think the most effective COVID-19 vaccine available reduces the chance of someone becoming ill with the disease? | [Slider]: Doesn't make any difference (0%) - - Stops all COVID-19 disease (100%). |
| Age | What is your age (in years) | [Numeric text entry] |
| Gender | What is your gender | Male<br>Female<br>Other<br>Prefer not to say |
| Ethnicity | How would you describe your ethnicity? | White: English / Welsh / Scottish / Northern Irish / British<br>White: Irish<br>White: Gypsy or Irish Traveller<br>Any other White |

| | | |
|---|---|---|
| | | background<br>Mixed: White and Black Caribbean<br>Mixed: White and Black African<br>Mixed: White and Asian<br>Any other Mixed / Multiple ethnic background<br>Asian / Asian British: Indian<br>Asian / Asian British: Pakistani<br>Asian / Asian British: Bangladeshi<br>Asian / Asian British: Chinese<br>Any other Asian background<br>Black / Black British: Caribbean<br>Black / Black British: African<br>Any other Black / African / Caribbean background<br>Multiple ethnic background<br>Arab<br>Any other ethnic group |

*Denotes reverse score item. Items marked (O) were items from the original Freeman et al. (2020) scales which were excluded. Items marked (A) were additional items added to adapted scales.

#### *Adaptions to COVID-19 Vaccination Beliefs Scale*

As noted in the main text, we made several minor changes to the subscales outlined in Freeman et al. (2020). Specifically, in the main text analyses we removed one item from each of the perceived efficacy and concern over speed scales. These items were identified as loading weakly on the proposed factors in Freeman et al. (2020; supplementary material). We added two further face-valid items to each of these scales, and one further item to the concern over side effects scale (items noted in table S2.1) . Table S2.2 details the Cronbach’s alpha values for the original and adapted scales in our study, and the results of one-way ANOVAs examining the main effect of experimental condition on the original scale scores.

Table S2.2
Reliability of original and adapted scales and ANOVA results using original scales

| Scale | Reliability (α) | | ANOVA result (original scale) |
|---|---|---|---|
| | Original | Adapted | |

| | | | |
|---|---|---|---|
| Perceived efficacy | .59 | .87 | $F(4, 2091) = 0.49$, $p = .74$ |
| Side effect concern | .70 | .76 | $F(4, 2090) = 1.53$, $p = .19$ |
| Concern over speed | .79 | .79 | $F(4, 2091) = 1.80$, $p = .13$ |

**Appendix 3: Study 1 descriptive statistics and additional analyses.**

Table S3.1
Descriptive statistics for Study 1 primary outcomes

| Condition | Perceived efficacy | | Concern over side effects | | Concern over approval speed | | Vaccine intention | |
|---|---|---|---|---|---|---|---|---|
| | *M* | *SD* | *M* | *SD* | *M* | *SD* | *M* | *SD* |
| Control | 4.01 | 0.74 | 2.23 | 0.73 | 2.76 | 0.84 | 4.26 | 0.94 |
| Factbox | 4.04 | 0.69 | 2.33 | 0.76 | 2.81 | 0.83 | 4.21 | 0.96 |
| Q&A | 4.07 | 0.71 | 2.28 | 0.75 | 2.78 | 0.85 | 4.20 | 0.98 |
| Approval | 4.03 | 0.67 | 2.22 | 0.79 | 2.80 | 0.82 | 4.27 | 0.91 |
| Mechanism | 4.00 | 0.79 | 2.29 | 0.85 | 2.85 | 0.88 | 4.17 | 1.04 |

Table S3.2
Descriptive statistics for Study 2 secondary outcomes

| Condition | Estimated mild-moderate side effect frequency[a] | | Estimated severe side effect frequency[a] | | Estimated efficacy (%) | | Informed | | Decision certainty | |
|---|---|---|---|---|---|---|---|---|---|---|
| | *M* | *SD* | *M* | *SD* | *M* | *SD* | *M* | *SD* | *M* | *SD* |
| Control | 1782.92 | 2515.07 | 540.97 | 1500.58 | 75.44 | 20.28 | 3.94 | 0.91 | 4.20 | 1.02 |
| Factbox | 2663.86 | 2922.02 | 390.78 | 1038.12 | 75.78 | 19.93 | 4.21 | 0.84 | 4.28 | 0.94 |
| Q&A | 1518.20 | 2313.52 | 466.17 | 1420.21 | 78.18 | 20.62 | 4.28 | 0.86 | 4.27 | 1.01 |
| Approval | 1703.54 | 2638.13 | 497.99 | 1435.16 | 75.79 | 20.74 | 4.05 | 0.86 | 4.20 | 0.98 |
| Mechanism | 1919.73 | 2728.63 | 581.94 | 1616.93 | 74.53 | 21.97 | 4.04 | 0.89 | 4.23 | 0.97 |

[a]per 10,000 people vaccinated

***Analysis of Study 1 secondary 'message only' outcomes***

Here we report the results of analyses investigating the effect of experimental condition on participants' perceptions of the information presented. These include measures of how trustworthy, engaging, easy to understand, and believable participants found the information, as well as the level of effort required to read the message, and the perceived quality of the evidence underlying the information. As these items were only shown to participants who read a message, the control group is excluded.

Means and standard deviations are reported in Table S3.3 and the results of one-way ANOVAs comparing the four groups are reported in table S3.4. Means are plotted in Figure S3.1, with significant post-hoc pairwise differences noted (Tukey's HSD).

Table S3.3
Means and standard deviation for secondary, information-only variables.

| Condition | Trust in info | | Engagement | | Understood | | Effort | | Believable | | Quality of evidence | |
|---|---|---|---|---|---|---|---|---|---|---|---|---|
| | *M* | *SD* | *M* | *SD* | *M* | *SD* | *M* | *SD* | *M* | *SD* | *M* | *SD* |
| Factbox | 5.20 | 1.26 | 5.34 | 1.34 | 5.71 | 1.25 | 4.37 | 1.75 | 5.32 | 1.32 | 5.17 | 1.32 |
| Q&A | 5.54 | 1.33 | 5.68 | 1.28 | 6.03 | 1.09 | 4.35 | 1.81 | 5.62 | 1.37 | 5.48 | 1.37 |
| Approval | 5.58 | 1.16 | 5.17 | 1.34 | 5.32 | 1.25 | 4.81 | 1.56 | 5.55 | 1.21 | 5.53 | 1.17 |
| Mechanism | 5.61 | 1.21 | 5.44 | 1.36 | 5.44 | 1.35 | 4.86 | 1.64 | 5.63 | 1.28 | 5.54 | 1.27 |

Table S3.4
Results of one-way ANOVA tests

| Variable | *F* statistic |
|---|---|
| Trust in info | $F(3, 1680) = 9.51$, $p < .001$, $\eta^2 = 0.02$ |
| Engagement | $F(3, 1681) = 10.98$, $p < .001$, $\eta^2 = 0.02$ |
| Understood | $F(3, 1681) = 27.50$, $p < .001$, $\eta^2 = 0.05$ |
| Effort | $F(3, 1681) = 10.98$, $p < .001$, $\eta^2 = 0.02$ |
| Believable | $F(3, 1680) = 5.23$, $p < .01$, $\eta^2 = 0.01$ |
| Quality of evidence | $F(3, 1680) = 7.50$, $p < .001$, $\eta^2 = 0.01$ |

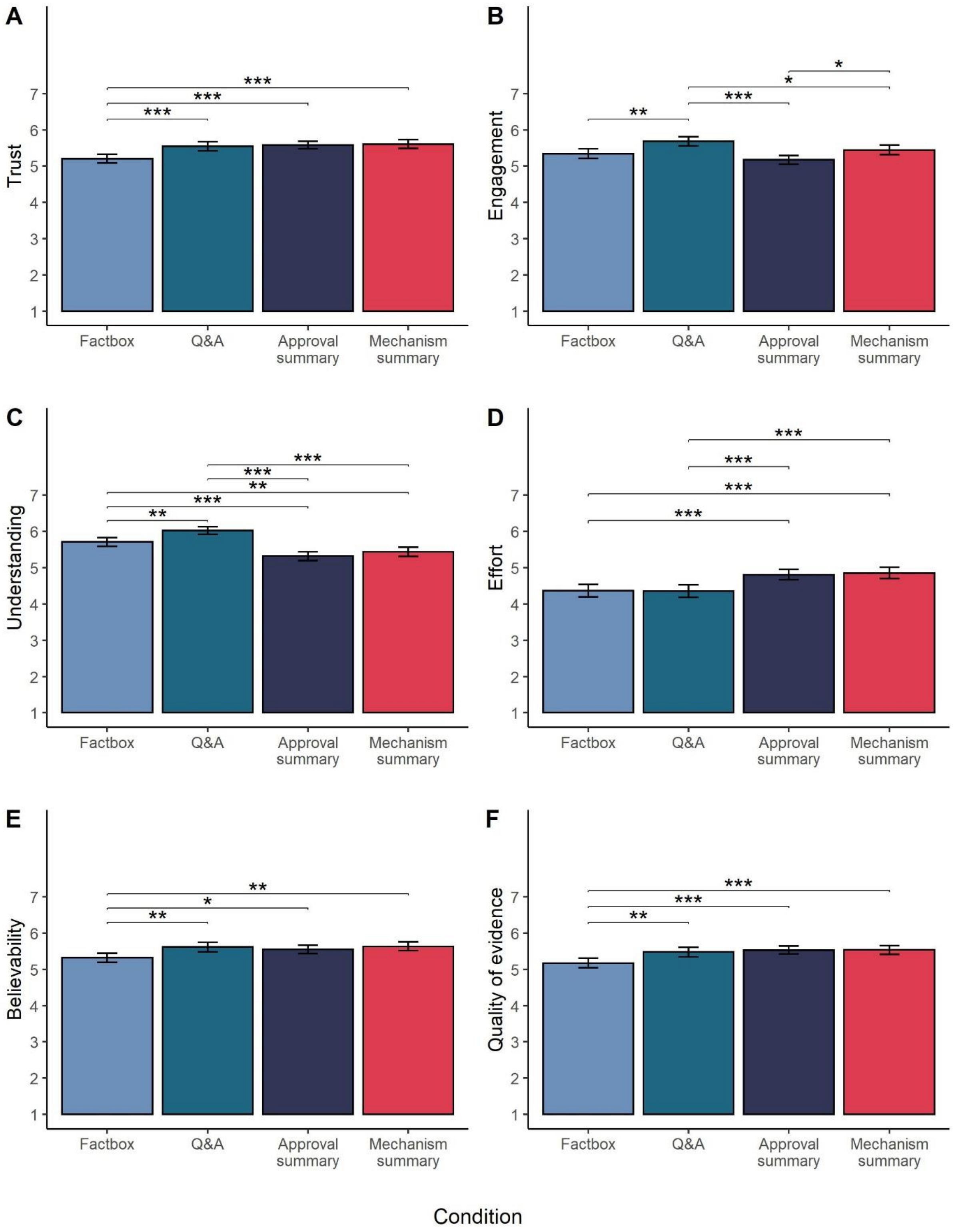


Figure S3.1: Mean (95%CI) levels of (A) trust in information presented, (B) rating of engagement, (C) ease of understanding, (D) effort required to read, (E) believability, (F) perceived quality of evidence underlying information. Horizontal bars and asterisks indicate significant pairwise difference between conditions, based on post-hoc tests (Tukey's HSD), *$p < .05$, **$p < .01$, ***$p < .001$.

**Appendix 4. Study 2 messages**

Table S4.1. Study 2 Message content

| Message length | Message Content | | |
|---|---|---|---|
| | No Caution | Medium Caution | High Caution |
| Short | **The COVID-19 vaccines have been shown to be safe and effective.**<br>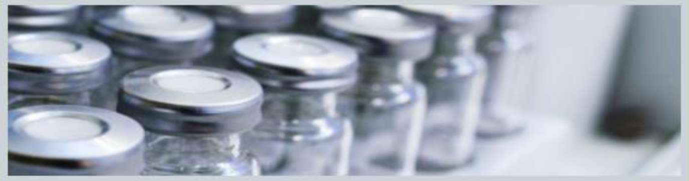 | **The COVID-19 vaccines are up to 95% effective - not 100%.**<br><br>**So, keep remembering: Hands, Face, Space.**<br>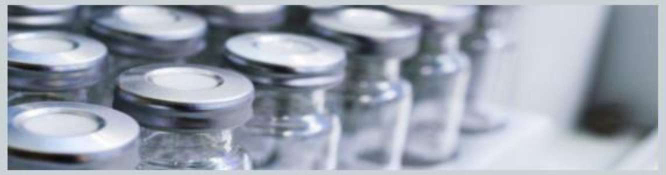 | **The COVID-19 vaccines don't give you 100% protection and you can still pass the disease to others.**<br><br>**Keep remembering: Hands, Face, Space.**<br>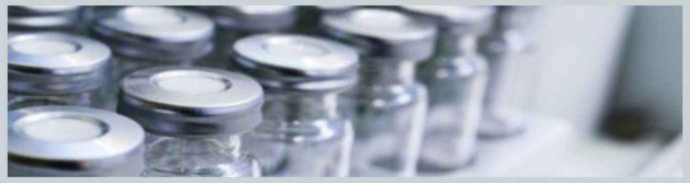 |
| Long | The COVID-19 vaccines available in the UK have been shown in clinical trials to be safe and effective.<br><br>They have been tested in over 20,000 adults, covering a wide range of ages and ethnicities and including people with underlying health conditions.<br><br>They have been shown to reduce the chance of developing serious COVID-19 symptoms by up to 95% and be very safe. The vaccine cannot give you COVID-19 infection.<br><br>The vaccine is an important part of our way out of the pandemic, and is being offered free to all on the NHS. Please take your slot when it is offered to you. Our NHS staff and volunteers are working long hours to ensure that everyone can get their vaccine safely and as quickly as possible.<br><br>For current guidance, see www.gov.uk/ coronavirus<br>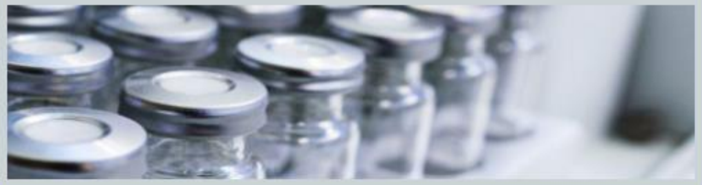 | The COVID-19 vaccines available in the UK have been shown in clinical trials to be safe and effective. The vaccine cannot give you COVID-19 infection, and a full course will reduce your chance of becoming seriously ill.<br><br>We do not yet know whether it will stop you from catching and passing on the virus, but we do expect it to reduce this risk.<br><br>So, it is still important to follow the guidance in your local area to protect those around you.<br><br>To protect yourself and your family, friends and colleagues you still need to:<br>• practise social distancing<br>• wear a face mask<br>• wash your hands carefully and frequently<br>• follow the current guidance at www.gov.uk/ coronavirus<br>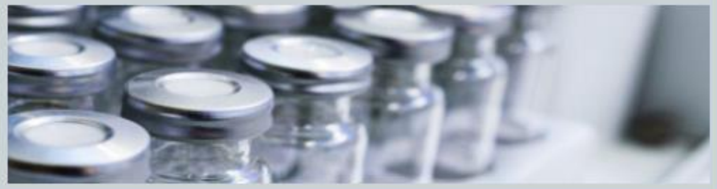 | The COVID-19 vaccines available in the UK have been shown to reduce the chance of developing serious COVID-19 symptoms by between 41-98%.<br><br>No vaccine gives you 100% protection.<br><br>They were tested in over 20,000 adults, covering a wide range of ages and ethnicities and including people with underlying health conditions. The vaccine cannot give you COVID-19 infection.<br><br>We do not yet know whether the vaccine will stop you from catching and passing on the virus, but we do expect it to reduce this risk.<br><br>So, to protect yourself and your family, friends and colleagues you still need to:<br>• practise social distancing<br>• wear a face mask<br>• wash your hands carefully and frequently<br>• follow the current guidance at www.gov.uk/ coronavirus<br>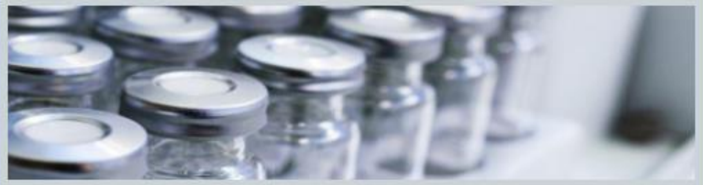 |

**Appendix 5: Study 2 survey items**

Measures of vaccine intentions, perceived efficacy, estimated efficacy, and message ratings (trustworthiness, engagement, believability, understanding, effort required, and perceived quality of evidence) were identical to Study 1, see Table S2.1. Table S5.1 details the additional items included in Study 2.

Table S5.1
Additional Study 2 items

| Scale | Label | Responses |
|---|---|---|
| Intention to engage in protective behaviours post vaccination | Imagine that you had received a COVID-19 vaccination. After receiving the vaccine, how likely is it that you would engage in the following behaviours because of COVID-19? – Social distancing - staying more than 1m from people not in your bubble<br><br>Washing your hands carefully and frequently<br><br>Wearing a face mask in public spaces<br><br>Avoiding social gatherings<br><br>Working from home whenever possible<br><br>Avoiding public transport | Very unlikely (1)<br>Moderately unlikely (2)<br>Slightly unlikely (3)<br>Neither likely nor unlikely (4)<br>Slightly likely (5)<br>Moderately likely (6)<br>Very likely (7) |
| Intention to follow guidance once vaccinated | Imagine that there are still coronavirus rules or restrictions in place after you have had the coronavirus vaccine. How much do you agree or disagree with the following statement below?<br>"I would still follow whatever coronavirus rules or restrictions were in place as strictly as I was before getting a vaccine" | Strongly disagree (1)<br>Disagree (2)<br>Somewhat disagree (3)<br>Neither agree nor disagree (4)<br>Somewhat agree (5)<br>Agree (6)<br>Strongly agree (7) |
| Worry over risk behaviours if vaccinated | If you had received the COVID-19 vaccine, how<br>worried would you feel doing each of the<br>*following?* - Shopping in a busy supermarket<br><br>Eating indoors in a restaurant with a small group of friends | Not at all worried (1)<br>(2)<br>(3)<br>(4)<br>(5)<br>(6)<br>Very worried (7) |

| | | |
|---|---|---|
| | Drinking in a pub garden with a small group of friends<br><br>Going to a large cinema<br><br>Visiting an elderly person in a nursing home<br><br>Attending Accident and Emergency in a city hospital | |
| Emotional response: enthusiasm | How did the vaccine information we showed you earlier make you feel? For each of the feeling below, please indicate on the sliding scales (from 'Not at all' to 'Extremely') – Enthusiastic<br><br>Proud<br><br>Hopeful | Sliding scale from 'Not at all' (0) to Extremely (100) |
| Emotional response: anxiety | Scared<br><br>Worried<br><br>Afraid | |
| Emotional response: aversion | Resentful<br><br>Bitter<br><br>Angry<br><br>Hateful | |
| Perceived public importance of vaccines | If I get the COVID-19 vaccine it will be: | Really helpful for the community around me (1)<br>Helpful for the community around me (2)<br>Neither helpful nor unhelpful for the community around me (3)<br>Unhelpful for the community around me (4)<br>Really unhelpful for the community around me (5) |
| | If individuals like me get the COVID-19 vaccine it will: | Save a large number of lives (1)<br>Save some lives (2)<br>Have no impact (3)<br>Lead to more deaths (4)<br>Lead to a large number of deaths (5) |

| | | |
|---|---|---|
| | If many people do not get the vaccine this: | Will be dangerous (1)<br>May be dangerous (2)<br>Will have no consequences at all (3)<br>May be good (4)<br>Will be good (5) |
| | The COVID-19 vaccine will: | Greatly strengthen my immune system (1)<br>Strengthen my immune system (2)<br>It will neither strengthen nor weaken my immune system (3)<br>Weaken my immune system (4)<br>Greatly weaken my immune system (5) |
| | Taking the COVID-19 vaccine: | Will give me complete freedom to get on with life just as before (1)<br>Will give me greater freedom (2)<br>Will have no effect on my freedom (3)<br>Will restrict my freedom (4)<br>Will completely restrict my freedom to get on with life (5) |

**Appendix 6: Study 2 descriptive statistics and additional results**

Table S6.1
Primary outcome means and SDs across experimental conditions

| Length | Content | Vaccine intentions | | Follow guidance if vaccinated | | Engage in protective behaviour if vaccinated | |
|---|---|---|---|---|---|---|---|
| | | M | SD | M | SD | M | SD |
| Control | Control | 4.21 | 0.99 | 6.19 | 1.25 | 5.95 | 1.24 |
| Long | No Caution | 4.37 | 0.90 | 6.2 | 1.36 | 5.94 | 1.33 |
| Long | Medium Caution | 4.31 | 0.89 | 6.23 | 1.24 | 6.11 | 1.12 |
| Long | High Caution | 4.31 | 0.92 | 6.37 | 1.10 | 6.13 | 1.06 |
| Short | No Caution | 4.37 | 0.87 | 6.22 | 1.30 | 6.04 | 1.22 |
| Short | Medium Caution | 4.31 | 0.97 | 6.29 | 1.24 | 6.10 | 1.19 |
| Short | High Caution | 4.21 | 1.09 | 6.14 | 1.33 | 6.00 | 1.28 |

Table S6.2
Secondary outcome means and SDs across experimental conditions

| Length | Content | Worry over risk behaviours if vaccinated | | Perceived public importance of COVID-19 vaccines | | Perceived efficacy | | Estimated efficacy (%) | | Informed | | Certainty | |
|---|---|---|---|---|---|---|---|---|---|---|---|---|---|
| | | M | SD | M | SD | M | SD | M | SD | M | SD | M | SD |
| Control | Control | 4.54 | 1.73 | 4.07 | 0.66 | 3.89 | 0.77 | 70.99 | 23.07 | 4.01 | 0.93 | 4.13 | 1.04 |
| Long | No Caution | 4.38 | 1.71 | 4.16 | 0.59 | 4.14 | 0.7 | 78.65 | 20.76 | 4.20 | 0.83 | 4.30 | 0.95 |
| Long | Medium Caution | 4.71 | 1.72 | 4.08 | 0.61 | 3.93 | 0.71 | 73.75 | 19.97 | 4.22 | 0.81 | 4.27 | 0.86 |
| Long | High Caution | 4.71 | 1.61 | 4.05 | 0.61 | 3.92 | 0.73 | 74.65 | 20.27 | 4.09 | 0.93 | 4.19 | 1.02 |
| Short | No Caution | 4.50 | 1.74 | 4.15 | 0.59 | 4.04 | 0.72 | 76.10 | 20.00 | 4.20 | 0.92 | 4.29 | 0.97 |
| Short | Medium Caution | 4.60 | 1.71 | 4.12 | 0.62 | 4.03 | 0.73 | 78.69 | 20.95 | 4.12 | 0.90 | 4.21 | 1.03 |
| Short | High Caution | 4.69 | 1.77 | 4.02 | 0.73 | 3.86 | 0.83 | 74.25 | 21.79 | 4.06 | 0.92 | 4.24 | 0.96 |

#### ***Analysis of Study 2 secondary 'message only' outcomes***

Here we report the results of analyses investigating the effect of experimental condition on participants' perceptions of the information presented. These include measures of how trustworthy, engaging, easy to understand, and believable participants found the information, as well as the level of effort required to read the message, the perceived quality of the evidence underlying the information, and emotional response to the message. As these items were only shown to participants who read a message, the control group is excluded. As such we investigated the effects of message content (No Caution, Medium Caution, and High Caution) and length (Long, Short) using 2(type)x3(length) two way ANOVAs. Table S6.3 details the means and SDs for these outcomes across experimental groups.

Table S6.3
Means and SDs for secondary, information-only outcomes.

| Length | Content | Trust in info | | Engagement | | Emotion: enthusiasm | | Emotion: anxiety | | Emotion: Aversion | | Quality of evidence | | Believable | | Understood | | Effort | |
|---|---|---|---|---|---|---|---|---|---|---|---|---|---|---|---|---|---|---|---|
| | | M | SD | M | SD | M | SD | M | SD | M | SD | M | SD | M | SD | M | SD | M | SD |
| Long | No Caution | 5.71 | 1.36 | 5.76 | 1.23 | 67.22 | 26.18 | 24.36 | 28.57 | 14.36 | 23.78 | 5.52 | 1.45 | 5.66 | 1.39 | 6.29 | 0.99 | 3.58 | 2.02 |
| Long | Medium Caution | 5.65 | 1.28 | 5.69 | 1.20 | 64.36 | 25.12 | 29.55 | 26.68 | 17.76 | 24.53 | 5.42 | 1.28 | 5.57 | 1.29 | 6.26 | 0.94 | 3.77 | 1.92 |
| Long | High Caution | 5.65 | 1.35 | 5.64 | 1.28 | 63.34 | 25.72 | 29.72 | 27.90 | 18.02 | 24.29 | 5.4 | 1.44 | 5.51 | 1.37 | 6.27 | 0.89 | 4.04 | 2.00 |
| Short | No Caution | 5.68 | 1.47 | 5.56 | 1.29 | 66.96 | 27.33 | 21.39 | 24.65 | 13.42 | 22.34 | 5.37 | 1.59 | 5.61 | 1.43 | 6.28 | 0.97 | 3.63 | 2.15 |
| Short | Medium Caution | 5.46 | 1.42 | 5.47 | 1.30 | 62.23 | 25.87 | 26.70 | 25.04 | 16.12 | 22.91 | 5.33 | 1.43 | 5.41 | 1.45 | 6.16 | 1.13 | 3.83 | 2.05 |
| Short | High Caution | 5.51 | 1.45 | 5.51 | 1.30 | 59.69 | 27.79 | 30.20 | 25.59 | 18.15 | 22.31 | 5.29 | 1.49 | 5.49 | 1.46 | 6.19 | 1.00 | 3.57 | 1.95 |

We find there was no significant effect of either factor (or interaction between the two) for: perceived trustworthiness of the information; perceived quality evidence upon which the message was based; how believable the information was; how difficult the information was to understand .

We report a significant main effect of length (but not content) on how engaging participants found the message; Long messages, regardless of content, were rated as more engaging that Short messages $F(1, 1632) = 8.61$, $p < .01$, $\eta^2_G = 0.00$.

Considering participants' emotional response to the information, there were significant main effects of content (but not length) on the three emotion factors of Marcus et al.'s (Marcus et al., 2017) scale ($F$s(2, 1606-1611) 4.70-10.69, $p$s < .01, all $\eta^2_G = 0.01$). Pairwise comparison with Tukey's post hoc tests indicated that participants in the No Caution conditions reported greater enthusiasm ($M_{enthusiasm} = 67.09$, $SD = 26.57$) and less anxiety ($M_{anxiety} = 22.84$, $SD = 26.66$) than participants in the Medium ($M_{enthusiasm} = 63.31$, $SD = 25.49$, $p = 0.49$, $d = .14$; $M_{anxiety} = 28.15$, $SD = 25.91$, $p = .003$, $d = .20$) and High Caution conditions ($M_{enthusiasm} = 61.51$, $SD = 26.82$, $p = .001$, $d = .21$; $M_{anxiety} = 29.96$, $SD = 26.74$, $p < .001$, $d = .27$). Participants in the No Caution conditions also reported less aversion ($M = 13.89$, $SD = 23.05$) than those in the High Caution conditions ($M = 18.09$, $SD =23.30$, $p = .009$, $d = .18$).

We also report a significant interaction effect between length and content on reported effort required to interpret the message, $F(2, 1633) = 3.19$, $p = .04$, $\eta^2_G = 0.00$. This was followed up by separate one-way ANOVAs and post hoc tests examining the effect of content for Long ($F(2,813) = 3.72$, $p = .025$, $\eta^2_G = 0.01$), and Short messages ($F(2,820) = 1.22$, $p = .29$) separately. Results indicated that for Long messages, the High Caution message required significantly more effort ($M = 4.04$, $SD = 2.00$) than the No Caution message ($M = 3.58$, $SD = 2.02$, $p = .018$, $d = .2$.)
